# Cine phase contrast magnetic resonance imaging of calf muscle contraction in pediatric patients with cerebral palsy and healthy children: comparison of voluntary motion and electrically evoked motion

**DOI:** 10.1101/2023.08.02.23293313

**Authors:** Claudia Weidensteiner, Xeni Deligianni, Tanja Haas, Philipp Madoerin, Oliver Bieri, Meritxell Garcia, Jacqueline Romkes, Erich Rutz, Francesco Santini, Reinald Brunner

## Abstract

The aim of this study is to investigate the feasibility of phase contrast imaging for assessment of muscle function in children with cerebral palsy. Time-resolved cine phase contrast MRI at 3T was synchronized with (a) electrical muscle stimulation (EMS) of the calf muscle and (b) voluntary plantarflexion following visual instructions. Fourteen ambulatory pediatric patients with equinus and 13 normally developing, healthy children were scanned. Ten patients were scanned pre and post botulinum toxin treatment. Four patients and four healthy children performed voluntary plantarflexion additionally to EMS. The achieved force was higher for the voluntary task for both patients and healthy controls, but its periodicity was worse compared to the stimulated contraction in patients. Therefore, it was possible to acquire contraction velocity time courses showing two distinct velocity peaks – the first for voluntary muscle contraction and the second for release – in four out of four healthy controls but in none of the patients. During EMS, two distinct velocity peaks could be resolved if the tolerated current was high enough to evoke sufficient contraction. That was achieved in 21% of the scans in patients (15 out of 72 scans during EMS in total) compared to 82% (14 out of 17) in healthy children. Then, the data quality was sufficient to generate strain maps. However, it was not possible to detect an effect of botulinum toxin in these patients since we did not succeed in acquiring data with sufficient quality both pre- and post-treatment. In conclusion, both EMS and voluntary motion worked well in healthy, normally developing children. Compliance was higher for the voluntary task for both patients and healthy controls. In conclusion, it was necessary to use EMS for a successful measurement of contraction/release velocity and strain in CP patients and although in this cohort the results were inconclusive, in the future optimization of the stimulation protocol could increase the acceptance and improve the output.

## Introduction

Cerebral palsy (CP) is a sensorimotor dysfunction caused by damage to the developing brain leading to weakness and spasticity of the affected muscles (1). In patients with affected legs, contractures lead to toe-walking (equinus) and gait disorders. Botulinum toxin A (BTX) has been established as an important treatment modality, particularly for the management of spasticity (2) to reduce inadequate muscle activity in CP patients, e.g. the calf muscles. BTX is injected locally into the affected calf muscles to control functional equinus and spasticity and thus improve gait function. The motivation for this study in CP patients was to find a method to assess muscle function with the aim of measuring the extent of the paralyzing effect of BTX with MRI. Time-resolved cine phase contrast (PC) MRI can be used to study muscle function and was applied to assess voluntary leg muscle contraction in adults, e.g., as described by Mazzoli et al. and Sinha et al. (3–7). The cine PC approach relies on the consistently repeated execution of the motion task over some minutes until data acquisition is completed. During the acquisition time, phase maps at each time point of the periodic motion cycle are acquired with a time resolution of several milliseconds. From these phase maps, maps of velocity vectors can be calculated showing the motion of the muscle during the cycle. However, in CP, there is the challenge that patients cannot always freely control movement of their limbs and a regular periodic motion cannot be performed. Therefore, voluntary exercise paradigms might not be adequate for examination. A PC MRI method to measure involuntary muscle contraction induced by electrical muscle stimulation (EMS) has been developed (8) and applied in adult healthy volunteers and patients (9,10), but not in children, yet. EMS evokes a regular, controlled motion but the stimulation current may feel unpleasant, and the maximum tolerated current may be too low to evoke motion – especially in young children. We applied both methods in pediatric patients with CP and healthy children to check the feasibility of a children-adapted experimental set-up for assessing calf muscle function with time-resolved PC MRI.

## Materials and methods

Two groups of children were recruited between February 22nd, 2018 and July 29th, 2021 and included in the study: 13 healthy, normally developing controls (6 males and 7 females, age range at MRI scan 9 to 16 years) and 14 ambulatory patients diagnosed with CP (9 males and 3 females, age range 8 to 15 years, 4 bilateral spastic CP functionally diparetic and 10 unilateral spastic CP functionally hemiparetic) were prospectively monitored. Participant characteristics can be found in Tables 1 and 2. Results of the clinical examinations and the gait analysis are presented in (11). All patients were toe-walking and were scheduled for muscle lengthening surgery with a preoperative BTX test injection to investigate potential muscle weakening, which can be a side effect of the muscle lengthening surgery (12). Functional mobility level was classified as Gross Motor Functional Classification Scale level (GMFCS) I (n = 11), II (n = 2), and III (n=1) (13). Eleven of the 14 patients received BTX injections in the affected leg (dose see Table 1), as described in (14). In the bilaterally affected patients, only the more affected leg was treated. In the healthy controls, the dominant leg (defined as preferred leg for kicking a ball) was scanned. Healthy controls were scanned once or twice (reason described below), patients were scanned one to three times (pre-BTX, 6 weeks post-BTX, 12 weeks post BTX), see Tables 3 and 4. Exclusion criteria were previous surgeries on the affected limb(s), claustrophobia, and difficulties in following instructions in the scanner. The study was conducted in accordance with the Declaration of Helsinki. Prior to participation, informed written consent was obtained from the participants’ parents and additionally from those participants aged 12 years or older. The study was approved by the local ethics committee (Ethikkommission Nordwest- und Zentralschweiz, EKNZ BASEC 2016-01408).

**Table 1.** Participant characteristics of CP patients.

| patient number | sex | age range [years] | type of CP | GMFCS | height range [cm] | BMI percentile range [%] | BTX total dose [U] | treated muscles |
| --- | --- | --- | --- | --- | --- | --- | --- | --- |
| 1 | m | 12-16 | uni | I | 140-150 | 0-10 | 150 | Gl+Gm+S |
| 2 | m | 7-11 | uni | I | 150-160 | 90-100 | 150 | Gl+Gm+S |
| 3 | f | 7-11 | uni | I | 140-150 | 20-30 | 150 | Gl+Gm+S |
| 4 | m | 7-11 | bi | I | 130-140 | 40-50 | 150 | Gl+Gm+S |
| 5 | m | 12-16 | bi | II | 130-140 | 0-10 | 225 | Gl+Gm+S |
| 6 | m | 7-11 | bi | III | 120-130 | 0-10 | 200 | Gl+Gm |
| 7 | m | 7-11 | uni | II | 150-160 | 70-80 | 0 | – |
| 8 | m | 12-16 | uni | I | 160-170 | 60-70 | 0 | – |
| 9 | m | 7-11 | uni | I | 130-140 | 90-100 | 250 | Gl+Gm+S |
| 10 | m | 7-11 | uni | I | 130-140 | 0-10 | 0 | – |
| 11 | f | 7-11 | uni | I | 140-150 | 10-20 | 200 | Gl+Gm+S |
| 12 | m | 7-11 | uni | I | 120-130 | 50-60 | 150 | Gl+Gm |
| 13 | m | 12-16 | uni | I | 150-160 | 0-10 | 100 | Gl+Gm |
| 14 | f | 12-16 | bi | I | 160-170 | 50-60 | 300 | Gl+Gm+S |

11 of the 14 patients were treated with botulinum toxin A (BTX) injection in the calf muscles (gastrocnemius lateralis Gl, gastrocnemius medialis Gm, soleus S) of the (more) affected leg. CP: cerebral palsy, m: male, f: female, uni: unilateral, bi: bilateral, GMFCS: Gross Motor Functions Classification System, BMI: Body Mass Index, U: Units.

**Table 2.**
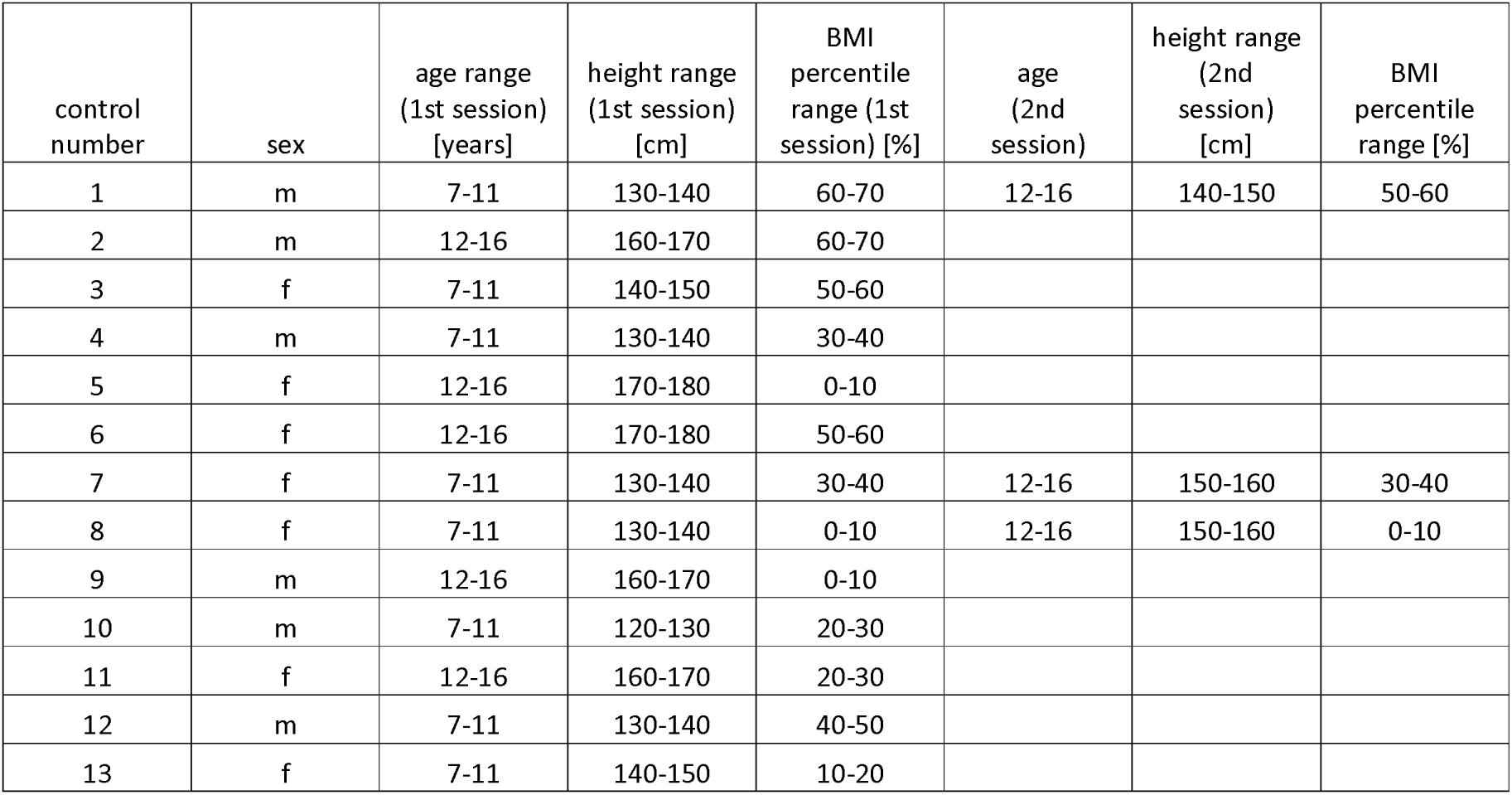
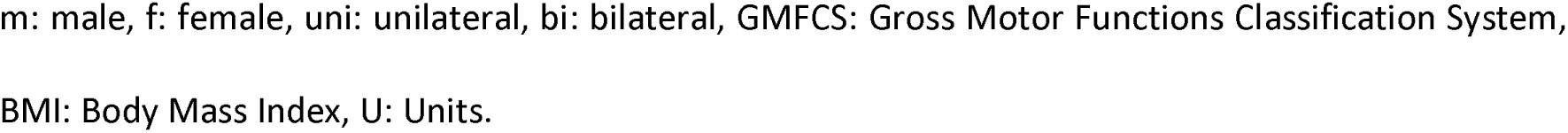
Participant characteristics of controls (healthy, normally developing children)

| control number | sex | age range (1st session) [years] | height range (1st session) [cm] | BMI percentile range (1st session) [%] | age (2nd session) | height range (2nd session) [cm] | BMI percentile range [%] |
| --- | --- | --- | --- | --- | --- | --- | --- |
| 1 | m | 7-11 | 130-140 | 60-70 | 12-16 | 140-150 | 50-60 |
| 2 | m | 12-16 | 160-170 | 60-70 |  |  |  |
| 3 | f | 7-11 | 140-150 | 50-60 |  |  |  |
| 4 | m | 7-11 | 130-140 | 30-40 |  |  |  |
| 5 | f | 12-16 | 170-180 | 0-10 |  |  |  |
| 6 | f | 12-16 | 170-180 | 50-60 |  |  |  |
| 7 | f | 7-11 | 130-140 | 30-40 | 12-16 | 150-160 | 30-40 |
| 8 | f | 7-11 | 130-140 | 0-10 | 12-16 | 150-160 | 0-10 |
| 9 | m | 12-16 | 160-170 | 0-10 |  |  |  |
| 10 | m | 7-11 | 120-130 | 20-30 |  |  |  |
| 11 | f | 12-16 | 160-170 | 20-30 |  |  |  |
| 12 | m | 7-11 | 130-140 | 40-50 |  |  |  |
| 13 | f | 7-11 | 140-150 | 10-20 |  |  |  |

m: male, f: female, uni: unilateral, bi: bilateral, GMFCS: Gross Motor Functions Classification System, BMI: Body Mass Index, U: Units.

**Table 3.** Overview of performed scans and data quality in patients.

| patient number | EMS |  |  |  |  |  |  |  | voluntary motion |  |  |
| --- | --- | --- | --- | --- | --- | --- | --- | --- | --- | --- | --- |
|  | pre BTX, VENC 25 | pre BTX, VENC 10 | 6w post BTX, VENC 25 | 6w post BTX, VENC 10 | 6w post BTX, non-treated leg, VENC 25 | 6w post BTX, non-treated leg, VENC 10 | 12w post BTX, VENC 25 | 12w post BTX, VENC 10 | pre BTX, VENC 10 | 6w post BTX, VENC 10 | 12w post BTX, VENC 10 |
| 1 | 15 mA, 5% |  | 14 mA, 4% | 14 mA, 1% | 13 mA, 5% | 13 mA, 0% | 18 mA, 3% | 18 mA, 2% |  |  |  |
| 2 | 16 mA, 21% |  | 15 mA, 9% | 15 mA, 9% | 15 mA, 3% | 15 mA, 2% | 16 mA, 2% | 16 mA, 2% |  |  |  |
| 3 | 16 mA, 2% |  | 15 mA, 3% |  | 17 mA, 1% |  | 19 mA, 13% | 19 mA, 3% |  |  |  |
| 4 | 15 mA, 14% | 15 mA, 14% | 18 mA, 23% |  | 19 mA, 1% |  | 19 mA, 9% | 19 mA, 7% |  |  |  |
| 5 | 14 mA, 10% | 14 mA, 10% | 9 mA, 15% | 9 mA, 13% | 7 mA, 16% | 7 mA, 15% | 16 mA, 2% | 16 mA, 2% |  |  |  |
| 6 | 8 mA, 2% | 8 mA, 2% | 15 mA, 3% | 15 mA, 1% |  |  | 8 mA, 0% | 10 mA, 3% |  |  |  |
| 7 | 10 mA, 3% | 20 mA, 2% |  |  |  |  |  |  |  |  |  |
| 8 | 10 mA, 1% |  |  |  |  |  |  |  |  |  |  |
| 9 | 14 mA, 8% | 14 mA, 8% | 15 mA, 1% | 15 mA, 0% | 16 mA, 0% | 18 mA, 0% | 13 mA, 2% | 13 mA, 2% |  |  | 10 N, 5% |
| 10 | 13 mA, 2% | 14 mA, 3% |  |  |  |  |  |  |  |  |  |
| 11 | 6 mA, 0% | 7 mA, 0% |  |  |  |  |  |  |  |  |  |
| 12 | 5 mA, 0% |  | 7 mA, 0% | 7 mA, 0% | 5 mA, 0% | 5 mA, 0% | 10 mA, 0% | 10 mA, 0% |  |  | 20 N, 11% |
| 13 | 12 mA, 0% |  | 19 mA, 2% | 19 mA, 1% |  |  | 12 mA, 0% | 14 mA, 0% |  | 20 N, 10% | 20 N, 21% |
| 14 | 13 mA, 11% | 13 mA, 10% | 8 mA, 0% | 8 mA, 1% | 8 mA, 0% | 8 mA, 0% |  |  | 20 N, 15% | 20 N, 33% |  |

The phase contrast cine scans were performed pre and post botulinum toxin A (BTX) injection with VENC values 10 or 25 cm/s and during stimulated motion (EMS) or a voluntary exercise task. Data quality was rated as good (*blue*), mediocre (*yellow*), and not sufficient (*red*). Empty cells mean the scans were not performed. The scans were performed in the treated leg. At 6w post BTX also the non-treated leg was scanned in some patients. The amplitude of the stimulation current (in mA) or the pre-set force level for the voluntary paradigm (in N) is also stated along with the achieved relative maximum force (in %). VENC: velocity encoding.

**Table 4.** Overview of performed scans and data quality in controls (healthy, normally developing children)

| control number | EMS |  |  | voluntary motion |  |
| --- | --- | --- | --- | --- | --- |
|  | 1st session, VENC 25 | 1st session, VENC 10 | 2nd session, VENC 10 | 1st session, VENC 10 | 2nd session, VENC 10 |
| 1 | 20 mA, 1% |  | 25 mA, 4% |  | 30 N, 10% |
| 2 | 23 mA, 1% |  |  |  |  |
| 3 | 24 mA, 0% |  |  |  |  |
| 4 | 26 mA, 15% |  |  |  |  |
| 5 | 28 mA, 9% |  |  |  |  |
| 6 | 22 mA, 8% |  |  |  |  |
| 7 | 20 mA, 14% |  | 23 mA, 23% |  | 40 N, 14% |
| 8 | 20 mA, 9% |  | 26 mA, 39% |  | 40 N, 16% |
| 9 | 16 mA, 15% |  |  |  |  |
| 10 | 23 mA, 5% |  |  |  |  |
| 11 | 21 mA, 16% |  |  |  |  |
| 12 | 13 mA, 3% | 13 mA, 3% |  |  |  |
| 13 |  | 22 mA, 19% |  | 40 N, 12% |  |

The phase contrast cine scans were performed in the dominant leg with velocity encoding (VENC) values 10 or 25 cm/s and during stimulated motion (EMS) or a voluntary exercise task. Data quality was rated as good (*blue*) and not sufficient (*red*). *Empty cells* mean the scans were not performed. The amplitude of the stimulation current (in mA) or the pre-set force level for the voluntary paradigm (in N) is also stated along with the achieved relative maximum force (in %).

The triceps surae muscle group of the (more) affected leg (the dominant leg of the healthy controls) was imaged in a 3T whole body clinical MRI scanner (Prisma, Siemens Healthineers, Erlangen, Germany). Two self-adhesive electrodes with MRI-visible markers were attached to the muscle belly. The children were placed feet first supine on the scanner bed and a flexible 18-channel body coil (Siemens Healthineers, Erlangen, Germany) was placed on the lower legs. The foot was attached with straps to a home-built foot pedal device with a built-in force sensor (15). Before starting the scans, the children were asked to press their front foot on the pedal with maximum force. The maximum voluntary force (MVF) was measured as the mean of two trials. One to three PC MRI experiments (evoked motion – using up to two PC MRI protocols with different velocity encoding VENC – and voluntary motion, see below) with periodic plantarflexion were performed in one session in each participant, as listed in Tables 3 and 4. The number of experiments was dependent on the compliance of the participants. We added a sequence with VENC 10 cm/s to the protocol during the course of the study, as we noted that a higher sensitivity compared to VENC 25 cm/s was needed (see Discussion). The voluntary paradigm was performed in four patients (in two of these patients at two timepoints) and in four healthy controls (Tables 3 and 4). In three of these controls, this scan session took place three years after the first MRI scan with evoked motion only, so also the evoked motion protocol with VENC 10 cm/s was repeated at the second MRI session. Finally, in eight patients the evoked motion was also measured in the contralateral, non-treated leg 6 weeks post BTX-injection.

The set-up for the EMS scans was similar to the one described in (8). For the induction of a periodic contraction, a two-channel commercially available EMS device (InTENSity Twin Stim III TENS and EMS Combo, Current Solutions LLC, Austin, TX) connected to a 5.1 x 8.9 cm^²^ rectangular self-adhesive gel-based electrodes (TENSUnits.com, USA) was used. For localization purposes, glycerin markers were placed on the surface of both electrodes. The electrodes were positioned over the gastrocnemii (approx. 7 cm below the popliteal fossa) and soleus (approx. 10 cm below the first electrode) muscles. The EMS cycle (1 s ramp up, 1 s plateau, 1 s ramp down, 1 s recovery, Fig 1 top left) had 30 pulses/s and a pulse width of 300 µs. The evoked force was recorded during the experiment with the force sensor in the pedal. Before starting the scan, the amplitude of the stimulation current was individually adjusted within the comfort levels (healthy controls 13–28 mA, patients 5–20 mA) to a point that an induced visible force output of at least 5% MVF was reached if possible. Two three-directional cine gradient echo PC velocity encoding sequences were applied using the same set-up. Both sequences were synchronized to the EMS-cycle. The acquisition window was 3.5 s during each EMS-cycle of 4 s. The sequence with VENC 10 cm/s was acquired with following parameters: 1 parasagittal slice based on the position of the two markers, voxels: 2.2×2.2×5.0 mm^3^, echo time 9.7 ms, bandwidth/pixel 400 Hz/px, flip angle 10°, FOV 280×140 mm^2^, acquisition time 2:28 min, 67 temporal phases, temporal resolution of 52 ms. The sequence with VENC 25 cm/s was performed with the following parameters: 3 parasagittal slices with a spatial resolution of 2.2×2.2×5.0 mm^3^, echo time 7.2 ms, bandwidth/ pixel = 400 Hz/px, flip angle 10°, FOV 280×140 mm^2^, 3 k-space lines per segment, acquisition time 3 min, 27 temporal frames, temporal resolution of 126 ms.

**Fig 1.**
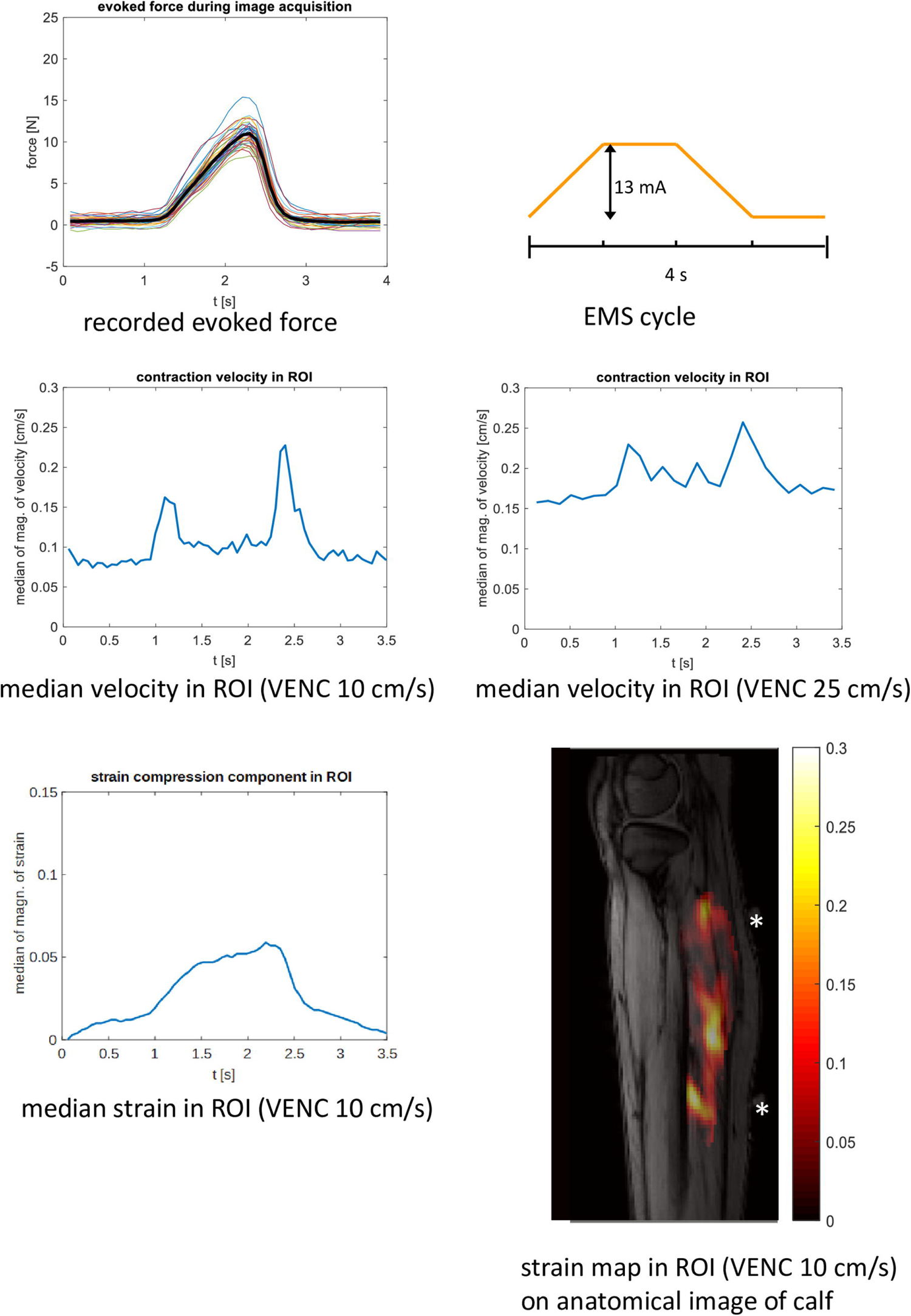
Cine phase contrast MR experiment with synchronized EMS in a girl with diparetic cerebral palsy. The experiments were performed in the more affected leg before BTX treatment. Top left: time courses of the evoked force on the pedal with the mean force curve (over the whole experiment) in black. Top right: current amplitude of the EMS cycle. Middle left: time course of the velocity magnitude (ROI median) showing two peaks, one at contraction (left) and one at release (right) acquired with VENC 10 cm/s. Middle right: for comparison, time course of the velocity magnitude (ROI median) acquired with VENC 25 cm/s. Bottom left: Bottom right: time course of the strain magnitude (ROI median of the strain compression component, acquired with VENC 10 cm/s). Bottom right: strain map at the temporal frame when the maximum occurs overlaid on an anatomical image of the calf (magnitude of the strain compression component, VENC 10 cm/s). The positions of the electrodes for stimulation are marked with asterisks. EMS: electrical muscle stimulation, VENC: velocity encoding.

In a subgroup of patients and healthy controls, a voluntary exercise paradigm was projected on a screen in the scanner room after a short rest phase following the scans with evoked motion. The paradigm consisted of a periodic curve with a pre-set amplitude (depending on the individual MVF, range 10–20 N in patients, 30–40 N in healthy children) and a period of 4 s. The shape of the predefined curve was the average of the force time courses of two adult volunteers who previously performed a periodic exercise (pressing the pedal for 1 s after a trigger signal every 4 s) in the same set-up. The force generated by the plantarflexion on the pedal was recorded and overlayed in real-time on the predefined curve. The paradigm was presented in the form of a video game: the fish – the force on the pedal – is supposed to hit as many bubbles – the predefined curve – as possible, see Fig 2 top left. The patient was supposed to follow the predefined curve by pressing the pedal. After a training period of 5 min the voluntary motion was performed during the scan. The code for the display of the exercise paradigm and the real-time force values were written in Python 3 (code available on https://github.com/BAMMri). The cine PC sequence with VENC 10 cm/s was applied with the acquisition synchronized to the default periodic curve, i.e., the acquisition window (3.5 s) started every 4 s in the resting phase. The sequence with VENC 25 cm/s was not performed for the voluntary exercise paradigm (due to its lower sensitivity compared to VENC 10 cm/s, see Discussion).

**Fig 2.**
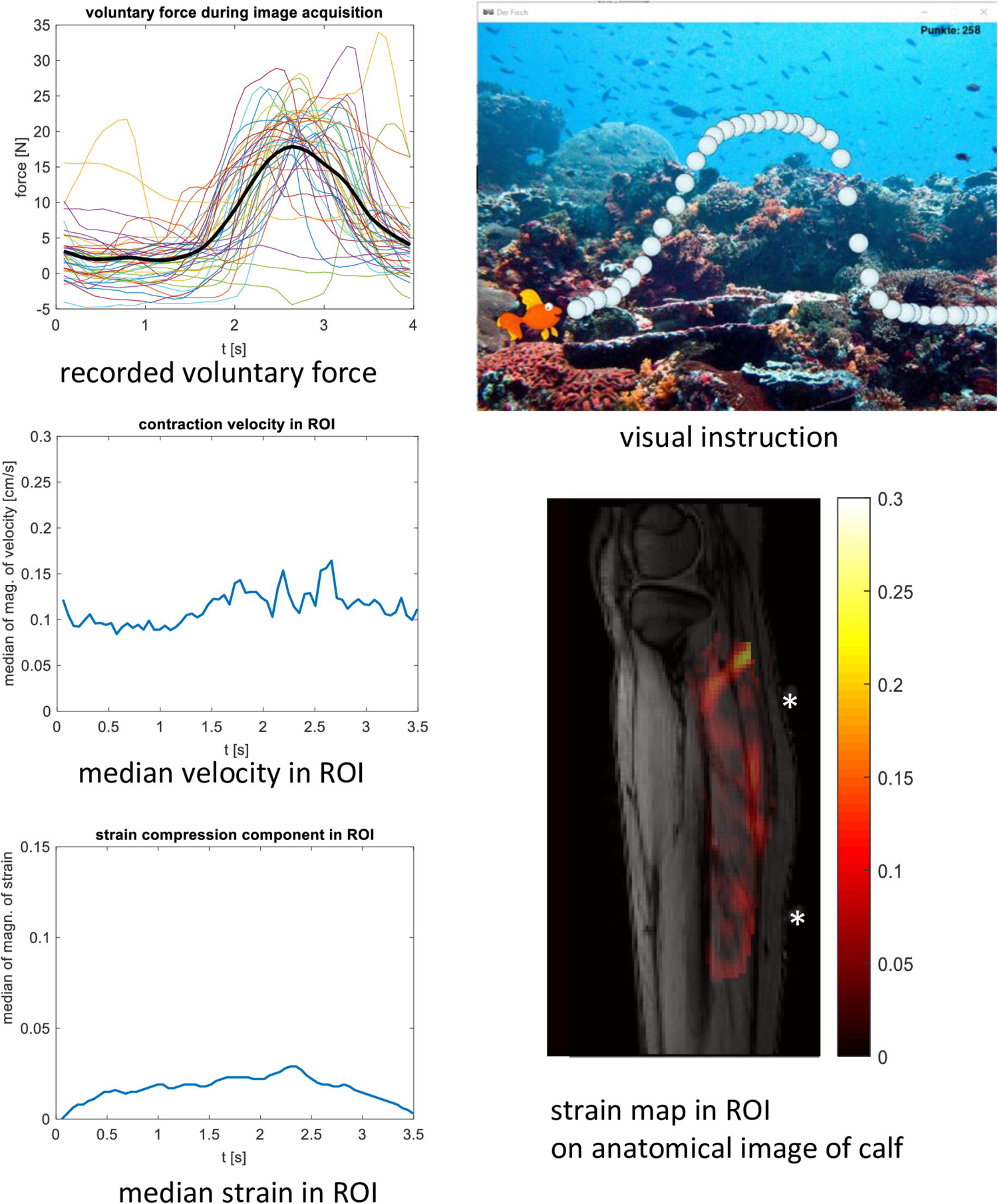
Cine phase contrast MR experiment with voluntary motion in a girl with diparetic cerebral palsy. The experiments were performed in the more affected leg before BTX treatment. The subject is the same as in Fig 1. Top left: time courses of the force on the pedal with the mean force curve in black. Top right: visual paradigm for instruction and feedback for the participant. Middle left: time course of the velocity magnitude (ROI median, acquired with VENC 10 cm/s) showing several small peaks. Bottom left: time course of the strain magnitude (ROI median of the strain compression component, acquired with VENC 10 cm/s). Bottom right: strain map at the temporal frame when the maximum occurs overlaid on an anatomical image of the calf (magnitude of the strain compression component). VENC: velocity encoding. BTX: botulinum toxin A.

Data were processed processing offline using Matlab (The Mathworks, Inc., Natick, MA, release R2017a). To visualize the contraction time course, a ROI was drawn manually on the triceps surae muscles visible in the image slice and the velocity vector field was displayed for every temporal phase (8). For the VENC 25 cm/s dataset, a ROI was drawn only on one slice, which was chosen after visual inspection (showing the largest area of triceps surae muscles). For the velocity time course, the ROI median of the magnitude of the velocity vectors was calculated for each phase. Strain vectors were extracted from the velocity and displacement fields as described in (8). The tensors were diagonalized, and the principal eigenvalue with the largest absolute value, which was the compression component, was visualized. For the strain time course, the ROI median of the magnitude of the strain compression component was calculated for each temporal frame. The data of control 3 was not processed as the MRI scan and force measurement under stimulation failed due to severe motion artifacts.

The velocity time courses were used to assess the data quality. The visibility of two clear velocity peaks (first peak for muscle contraction and one second later a peak for the release) was graded as good, the visibility of one peak or several not well-defined peaks was graded as mediocre, no visible peaks were graded as not sufficient.

Mean force time courses over the 4 s period were calculated from the recorded force data during each cine scan (i.e., as directly recorded with the foot pedal). The maximum force during stimulation was calculated as the difference between minimal and maximal value of the mean force time course. The relative maximum force was calculated as percentage of the MFV for each participant. If there was no detectable periodic motion in the force data, the maximum force was set to zero.

## Results

The maximum tolerated stimulation current and the developed force at this current showed a relatively large variability, even for the same participant between two sessions. The time course of the contraction speed showed two distinctive peaks for contraction and release (as in patient 14 shown in Fig 1) in 16 of 72 scans in patients and 14 of 17 scans in healthy children (as in control 13 shown in Fig 3). In these cases, the data quality was rated as good (Table 2), which was sufficient to calculate strain maps from the velocity vector. In these cases, the strain time courses showed a curved shaped like a bell (Figs 1 and 3). However, the strain maps (compression components) for the temporal frame where the maximum strain occurs were inhomogeneous with pronounced local differences (Figs 1 and 3). The maximum strain value in the time courses was in the range of 0.063 to 0.089 for the n=5 patient experiments with mediocre or good data quality pre BTX (acquired with VENC 25 cm/s), leading to a mean of 0.079 and a median of 0.081. In the normally developing children the corresponding range was 0.084 to 0.146 (n=9, acquired with VENC 25 cm/s), leading to a higher mean and median of 0.11 and 0.10, respectively. All the time courses for contraction/release velocity, strain, and evoked force during EMS can be found in S1 Graphs, and all the time courses for the direct comparison between EMS and voluntary motion paradigm can be found in S2 Graphs.

**Fig 3.**
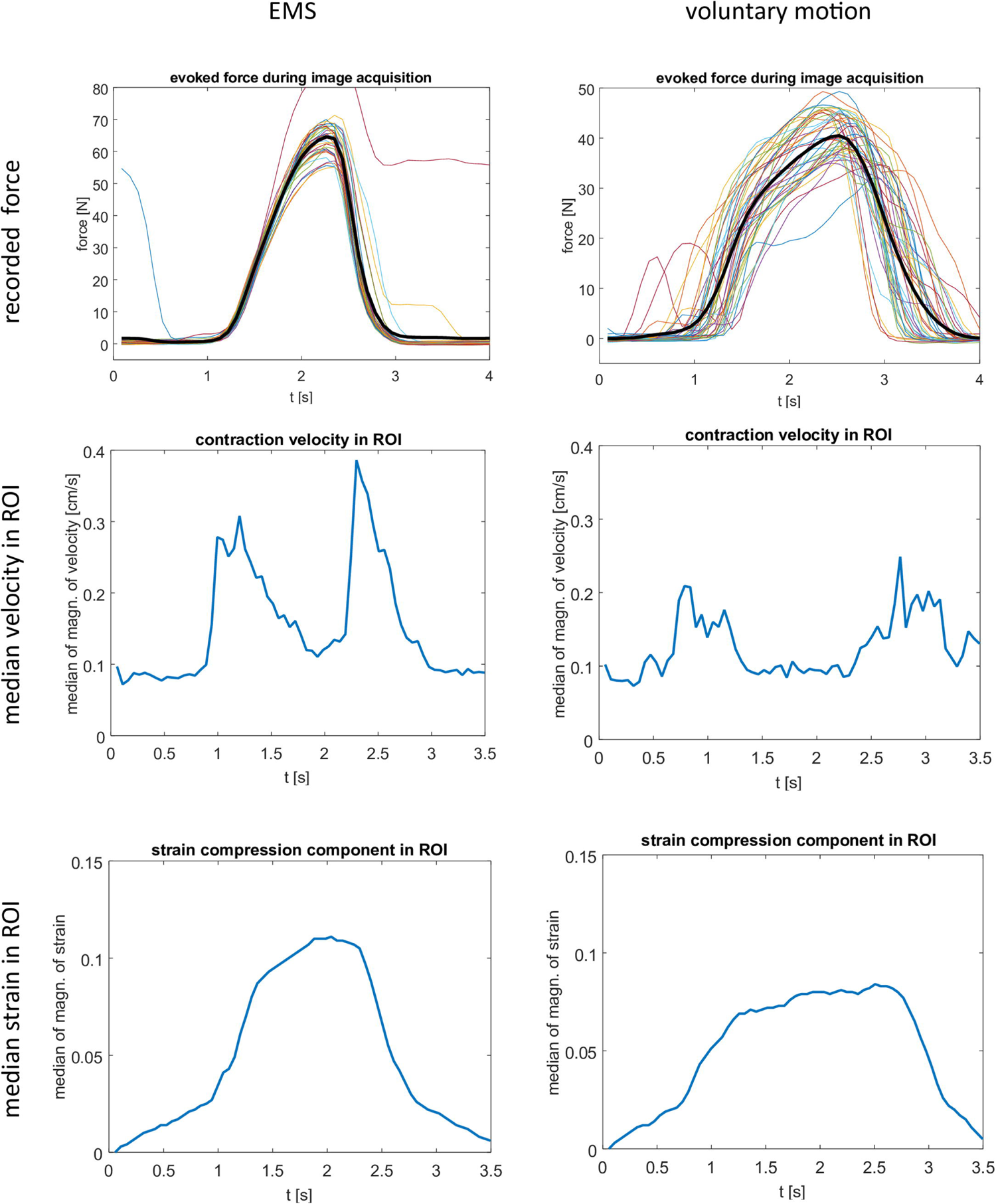
Cine phase contrast MR experiment with synchronized EMS and voluntary motion in a normally developing girl. The left column shows the results during stimulated motion (EMS), the right column during voluntary motion. Top row: time courses of the force on the pedal with the mean force curve in black for the stimulated and voluntary motion, respectively. Middle row: time course of the velocity magnitude (ROI median, acquired with VENC 10 cm/s) showing two peaks at contraction (left peak) and release (right peak). The peaks for the voluntary motion are relatively broad and not so well defined, corresponding to the lower periodicity of the force time courses. Bottom row: time courses of the strain compression component (ROI median, acquired with VENC 10 cm/s). EMS: electrical muscle stimulation, VENC: velocity encoding.

Contraction and release velocities were considerably below 1 cm/s. Due to the low contraction speeds, the sequence with the lower VENC of 10 cm/s, i.e., with the higher sensitivity for lower velocities, produced a higher signal-to-noise ratio, as can be seen in the velocity time courses (S1 Graphs). A higher noise level led to a higher offset in the plots with VENC 25 cm/s vs. 10 cm/s since the velocity time courses were calculated with absolute values. The data quality of VENC 25 cm/s and VENC 10 cm/s was directly compared in 32 cases. In 7 cases, the data quality was rated higher for VENC 10 cm/s, for the rest of the cases the data quality rating did not change.

Regarding the voluntary motion experiments, the pre-set amplitude of the voluntary force paradigm could be generated by the participants without much effort. The voluntary force amplitude was higher than the amplitude of the stimulated force (Table 2). However, the periodicity was lower, especially in patients (as shown in Fig 2 for patient 14, plots for all are shown in S2 Graphs). It was possible to resolve two distinct peaks in 4 of 4 scans in healthy children (i.e., data quality was rated as good, as shown in Fig 3 for control 13 and in S2 Graphs), and in 0 of 6 scans in patients (i.e., data quality was rated as mediocre – as in Fig 2 – or not sufficient, see Table 2).

The MVF (S3 Table) in the affected legs in patients before treatment was lower than in the legs of normally developing children (unpaired t-test, n=14/13, mean=119 N/247 N, median 113 N/260 N, p<0.001). The MFV in the less-affected legs (i.e., non-treated) in patients was not different to the one in legs of normally developing children (unpaired t-test, n=10/13, mean=181 N/247 N, median 157 N/260 N, p=0.09). The relative maximum force during stimulation was not different in patients both for affected and less-affected legs vs. healthy controls (unpaired t-test, n=14/13, mean=6%/9%, median=3%/9%, p=0.21 and n=8/13, mean=4%/9%, median 2%/9%, p=0.06).

In patients, there was no difference in relative maximum force of the treated leg pre vs. 6 weeks post BTX-injection (paired t-test, n=10, mean=7%/6%, median=7%/3%, p= 0.49) and pre vs. 12 w post BTX (paired t-test, n=9, mean=7%/3%, median=5%/2%, p=0.23). There was also no difference in the relative maximum force of the treated vs. the non-treated leg at 6 weeks post BTX-injection (paired t-test, n=8, mean=7%/4%, median=3%/2%, p=0.31). The MFV in the treated leg did not significantly differ pre vs 6 w or 12 w post BTX (paired t-test, n=10, mean=118 N/110 N, median=113 N/108 N, p=0.66; paired t-test, n=9, mean=119 N/135 N, median=121 N/113 N, p=0.23). The MFV in the treated leg was lower than in the non-treated leg at 6 w post BTX (paired t-test, n=10, mean=110 N/181 N, median=108 N/157 N, p= 0.006). There was no clear difference in data quality of the treated leg compared to the non-treated leg at 6 w post BTX.

## Discussion

The cine PC approach relies on the consistently repeated execution of the motion task over some minutes, which is problematic in patients with poor coordination skills and weakness, as in CP patients.

In this pilot study, the voluntary motion using our paradigm was well tolerated in both patients and healthy controls, and the children were very motivated. The force output during voluntary motion was larger than the one achieved with the maximum tolerated stimulation current, which is promising for detectable contraction velocities. Yet in patients during voluntary motion, the periodicity over the acquisition time was not sufficient to resolve two distinct velocity peaks for contraction and release. A periodic motion can be achieved with the EMS-paradigm. However, if the tolerated stimulation current is too low to evoke a detectable periodic motion, then the dynamic MR measurement fails. In our experience, this is the case in about 60% of the sessions with pediatric patients, where no velocity peak at all was visible in the velocity time courses (data quality rated as not sufficient).

In adults, the compliance and the tolerated stimulation current is higher, as demonstrated in (8–10,16). In these studies, two distinct velocity peaks could be detected, and the data quality was high enough to generate strain maps.

In healthy children both EMS and voluntary tasks worked well. In general, the relative maximum force on the pedal was higher than in the CP patients, which led to a detectable motion. The data quality was sufficient to generate meaningful velocity and strain time courses. Yet the strain maps were relatively inhomogeneous in the contracting muscles and conclusions about regional differences in the strain are difficult to be drawn. This inhomogeneous pattern in the strain maps has also been observed in healthy adults (8,9) in experiments with mediocre data quality (when there were no clear two velocity peaks present). The observed values for the maximum strain (in experiments with good or mediocre data quality) were in the same range as the strain values for adults in the calf during stimulated plantarflexion (9). The strain seemed to be lower in patients. Comparing EMS and voluntary motion, the evoked strain seemed to be higher than the strain during voluntary motion, both in patients and controls. Due to the high number of experiments without sufficient data quality we refrain from drawing further conclusions about strain quantification.

Our aim was to find an experimental set-up with EMS to allow us to generate strain maps in CP patients pre and post BTX therapy to evaluate the spatial extent of the paralyzing effect of BTX. However, it was not possible to achieve sufficient data quality pre and post BTX in the same patient. A comparison of strain maps pre and post BTX was thus not possible. A limitation of this study was that the voluntary paradigm was only applied for a subgroup of participants. This decision was based on our assumption that CP patients would not be able to perform a periodic voluntary motion over some minutes, therefore only a limited number of children were asked to undergo this task for comparison. The test in the subgroup of patients confirmed these doubts and data quality was also not sufficient for the voluntary paradigm due to the lack of periodicity. The recordings of the force on the pedal clearly showed lack of periodicity both in the voluntary motion and extra voluntary movements during stimulation (see S1 Graphs and S2 Graphs). So, these recordings were very helpful to explain the data quality of the cine PC MRI scans. It was also very practical to adjust the stimulation current with the participant already in the exam position inside the scanner.

The contraction velocities were below 1 cm/s, for both healthy children and patients and for both evoked and voluntary motion (see S1 Graphs and S2 Graphs). This is lower than the peak contraction values for evoked motion in adults, which was in the range of 0.5 to 10 cm/s in the thigh, and where VENC values of 25 cm/s (thigh) and 30 cm/s (lower leg) were used (8–10,16). A PC sequence with a low VENC is therefore appropriate in children. Since this was established after the initial tests and the start of the study, a delayed decision was taken to add the sequence with VENC 10 cm/s, which created an inherent inhomogeneity in the VENC values of the study. All the scans with the voluntary motion (which took place in the third year of the study) were acquired with the VENC 10 cm/s sequence only. It offered a higher sensitivity in combination with a better temporal resolution, but only one slice could be acquired in two and a half minutes compared to three slices in three minutes with the VENC 25 cm/s sequence. To achieve a lower VENC a higher gradient strength or longer gradient duration is needed which changes the timing of the sequence. Thus, multislice acquisition is not feasible with a low VENC in a short acquisition time.

One way to achieve higher success rates for these acquisitions could be a reduction in scan time. Compressed sensing reconstruction has already demonstrated the ability to speed up CP contrast MRI in the lower leg to 40 s for one slice (17) and 1 min 18 s for three slices (18). Patients might be able to achieve a sufficient periodicity in the voluntary motion over such a short period. Compressed sensing can also enable three-dimensional (3D) coverage of the total muscle belly in a reasonable acquisition time (3,4). A longer training period than the 5 minutes used in our study might also help to increase the periodicity and therefore improve data quality. However, an increase of the duration of the plantar flexion exercise may entail fatigue of the patients and an irregularity of motion.

In the EMS experiments, different settings of the stimulation parameters might be better tolerated. We chose our setting according to our experiences in adults and used stimulation parameters that are close to the settings for EMS used in physiotherapy in CP patients (19). However, in our experience, the maximum tolerated current was different from session to session in the same participant and depended on the mood and physical condition of the child at the day of the exam. It is very important for the success of the MRI acquisition to provide a pleasant atmosphere and to entertain the children by showing a movie or by other distractions during the exam inside the scanner.

In conclusion, cine PC MRI in combination with EMS or a voluntary motion paradigm to study plantarflexion is challenging in pediatric CP patients. The voluntary task in the form of a video game was well received, however, CP patients are often unable to perform a consistent periodic motion for a couple of minutes during data acquisition. Therefore, data quality was too low. EMS was not well tolerated in a big part of the sessions making clinical studies with several time points (like in our case, pre and post BTX treatment) difficult. It does nevertheless make repeatable exercise in the scanner potentially more feasible since an external device controls the movement. We already saw that the force curve became more periodic in the force pedal recordings. So, the critical point remains to increase acceptance and improve the sensation of the stimulation. In normally developing children, cine PC MRI both with EMS and the voluntary motion paradigm worked well. Contraction velocity and strain time courses were successfully generated in most cases. Although this method is still limited due to the difficulty in acquiring results with sufficiently high velocity and strain in such a cohort, it has proven its feasibility. With appropriate adaptations in the EMS stimulation protocol, it could be a very promising tool for studying muscle dynamics in healthy children and pediatric patients with neuromuscular diseases which is worth further investigation.

## Supporting information

S1 Graphs

S2 Graphs

## Data Availability

All relevant data are within the manuscript and its Supporting Information files.

**S1. Overview of all acquired time courses of the experiments under EMS.** First column: velocity time course, second column: strain time course (magnitude of contraction component), third column: all force time courses during MRI acquisition (plantarflexion on foot pedal), fourth column: mean force time course. Plots of all experiments in n=14 pediatric cerebral palsy (CP) patients and n=13 controls (healthy, normally developing children) are shown. The patients were scanned pre, 6 weeks, and 12 weeks post BTX injection. The velocity data were acquired with the cine phase contrast MR sequences with VENC 25 cm/s and/or VENC 10 cm/s. EMS: electrical muscle stimulation, VENC: velocity encoding, BTX: botulinum toxin A.

**S2. Comparison of the results under EMS and voluntary motion.** All the force plots of the plantarflexion on the foot pedal, velocity time courses, and strain time courses of the cine phase contrast MR sequences (VENC 10 cm/s) for all the four pediatric cerebral palsy (CP) patients and the four controls (healthy, normally developing children) that performed the voluntary paradigm experiments are shown. The amplitude of the EMS current (in mA), the pre-set force amplitude of the voluntary motion paradigm (in N), and the mean force amplitudes (in N) are stated in the graphs. The mean force is shown in black. For the strain time courses the magnitude of the contraction component is shown. The patients were scanned pre, 6 weeks and 12 weeks post BTX injection. EMS: electrical muscle stimulation, vol: voluntary motion, VENC: velocity encoding, BTX: botulinum toxin A.

**S3. Maximum voluntary force for plantarflexion in pediatric CP patients and healthy, normally developing children (controls).** Patient force measurements were performed pre, 6 weeks and 12 weeks post botulinum toxin A (BTX) injection.

## Notes

### Competing Interest Statement

The authors have declared no competing interest.

### Funding Statement

This study was funded by the Swiss National Science Foundation (http://www.snf.ch/; grant number 173292).

### Author Declarations

The Ethikkommission Nordwest- und Zentralschweiz gave ethical approval for this work (EKNZ BASEC 2016-01408).

## References

1. Mathewson MA, Lieber RL. Pathophysiology of muscle contractures in cerebral palsy. Phys Med Rehabil Clin N Am. 2015 Feb;26(1):57–67.

2. Strobl W, Theologis T, Brunner R, Kocer S, Viehweger E, Pascual-Pascual I, et al. Best clinical practice in botulinum toxin treatment for children with cerebral palsy. Toxins. 2015 May 11;7(5):1629–48.

3. Mazzoli V, Schoormans J, Froeling M, Sprengers AM, Coolen BF, Verdonschot N, et al. Accelerated 4D self-gated MRI of tibiofemoral kinematics. NMR Biomed. 2017;30(11):e3791.

4. Mazzoli V, Gottwald LM, Peper ES, Froeling M, Coolen BF, Verdonschot N, et al. Accelerated 4D phase contrast MRI in skeletal muscle contraction. Magn Reson Med. 2018 Nov;80(5):1799–811.

5. Sinha U, Malis V, Csapo R, Moghadasi A, Kinugasa R, Sinha S. Age-related differences in strain rate tensor of the medial gastrocnemius muscle during passive plantarflexion and active isometric contraction using velocity encoded MR imaging: potential index of lateral force transmission. Magn Reson Med. 2015 May;73(5):1852–63.

6. Sinha U, Malis V, Csapo R, Narici M, Sinha S. Shear strain rate from phase contrast velocity encoded MRI: Application to study effects of aging in the medial gastrocnemius muscle. J Magn Reson Imaging JMRI. 2018 Nov;48(5):1351–7.

7. Hernandez R, Sinha U, Malis V, Cunnane B, Smitaman E, Sinha S. Strain and Strain Rate Tensor Mapping of Medial Gastrocnemius at Submaximal Isometric Contraction and Three Ankle Angles. Tomography. 2023 Apr 11;9(2):840–56.

8. Deligianni X, Pansini M, Garcia M, Hirschmann A, Schmidt-Trucksäss A, Bieri O, et al. Synchronous MRI of muscle motion induced by electrical stimulation. Magn Reson Med. 2017 Feb;77(2):664– 72.

9. Deligianni X, Hirschmann A, Place N, Bieri O, Santini F. Dynamic MRI of plantar flexion: A comprehensive repeatability study of electrical stimulation-gated muscle contraction standardized on evoked force. PloS One. 2020;15(11):e0241832.

10. Deligianni X, Santini F, Paoletti M, Solazzo F, Bergsland N, Savini G, et al. Dynamic magnetic resonance imaging of muscle contraction in facioscapulohumeral muscular dystrophy. Sci Rep. 2022 May 4;12(1):7250.

11. Brunner R, De Pieri E, Wyss C, Weidensteiner C, Bracht-Schweizer K, Romkes J, et al. The Non-Affected Muscle Volume Compensates for the Partial Loss of Strength after Injection of Botulinum Toxin A. Toxins. 2023 Apr 3;15(4):267.

12. Rutz E, Hofmann E, Brunner R. Preoperative botulinum toxin test injections before muscle lengthening in cerebral palsy. J Orthop Sci Off J Jpn Orthop Assoc. 2010 Sep;15(5):647–53.

13. Palisano RJ, Rosenbaum P, Bartlett D, Livingston MH. Content validity of the expanded and revised Gross Motor Function Classification System. Dev Med Child Neurol. 2008 Oct;50(10):744–50.

14. Weidensteiner C, Madoerin P, Deligianni X, Haas T, Bieri O, Akinci D’Antonoli T, et al. Quantification and Monitoring of the Effect of Botulinum Toxin A on Paretic Calf Muscles of Children With Cerebral Palsy With MRI: A Preliminary Study. Front Neurol. 2021;12:630435.

15. Santini F, Bieri O, Deligianni X. OpenForce MR: A low-cost open-source MR-compatible force sensor. Concepts Magn Reson Part B Magn Reson Eng. 2018;48B(4):e21404.

16. Deligianni X, Klenk C, Place N, Garcia M, Pansini M, Hirschmann A, et al. Dynamic MR imaging of the skeletal muscle in young and senior volunteers during synchronized minimal neuromuscular electrical stimulation. Magma N Y N. 2020 Jun;33(3):393–400.

17. Malis V, Sinha U, Sinha S. Compressed sensing velocity encoded phase contrast imaging: Monitoring skeletal muscle kinematics. Magn Reson Med. 2020;84(1):142–56.

18. Malis V, Sinha U, Sinha S. 3D Muscle Deformation Mapping at Submaximal Isometric Contractions: Applications to Aging Muscle. Front Physiol. 2020 Dec 3;11:600590.

19. Kerr C, McDowell B, McDonough S. Electrical stimulation in cerebral palsy: a review of effects on strength and motor function. Dev Med Child Neurol. 2004 Mar;46(3):205–13.

