## Supplementary material for "Cine phase contrast magnetic resonance imaging of calf muscle contraction in pediatric patients with cerebral palsy and healthy children: comparison of voluntary motion and electrically evoked motion": S1 Graphs

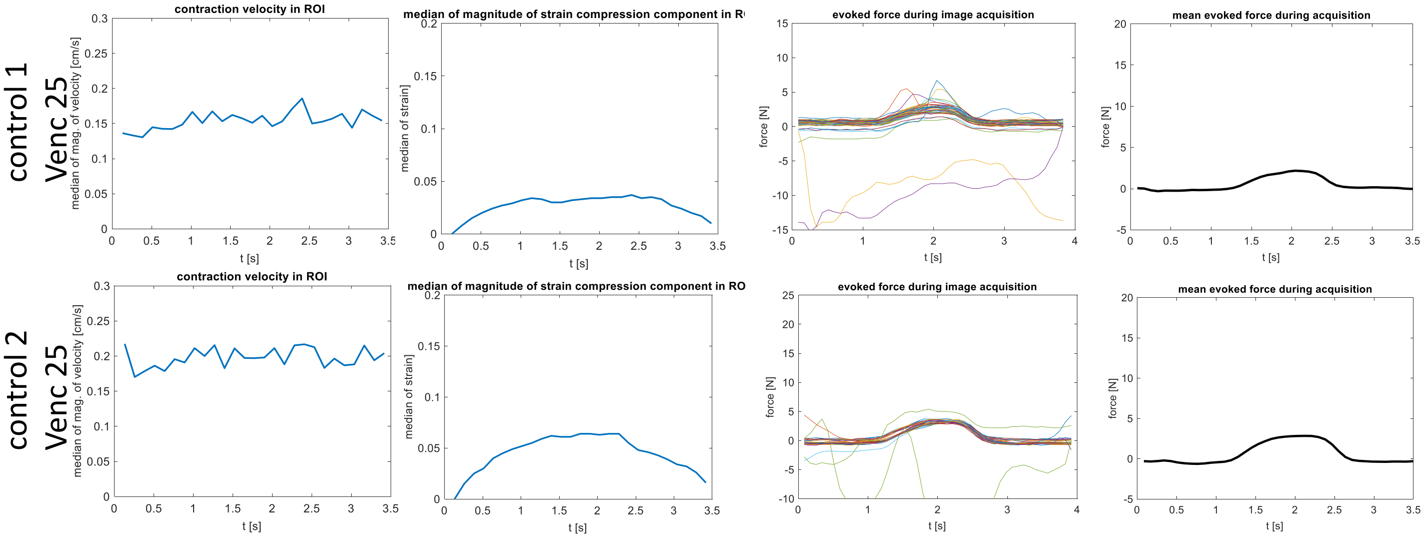


control 3

No data processing: experiment failed due to severe motion artifacts

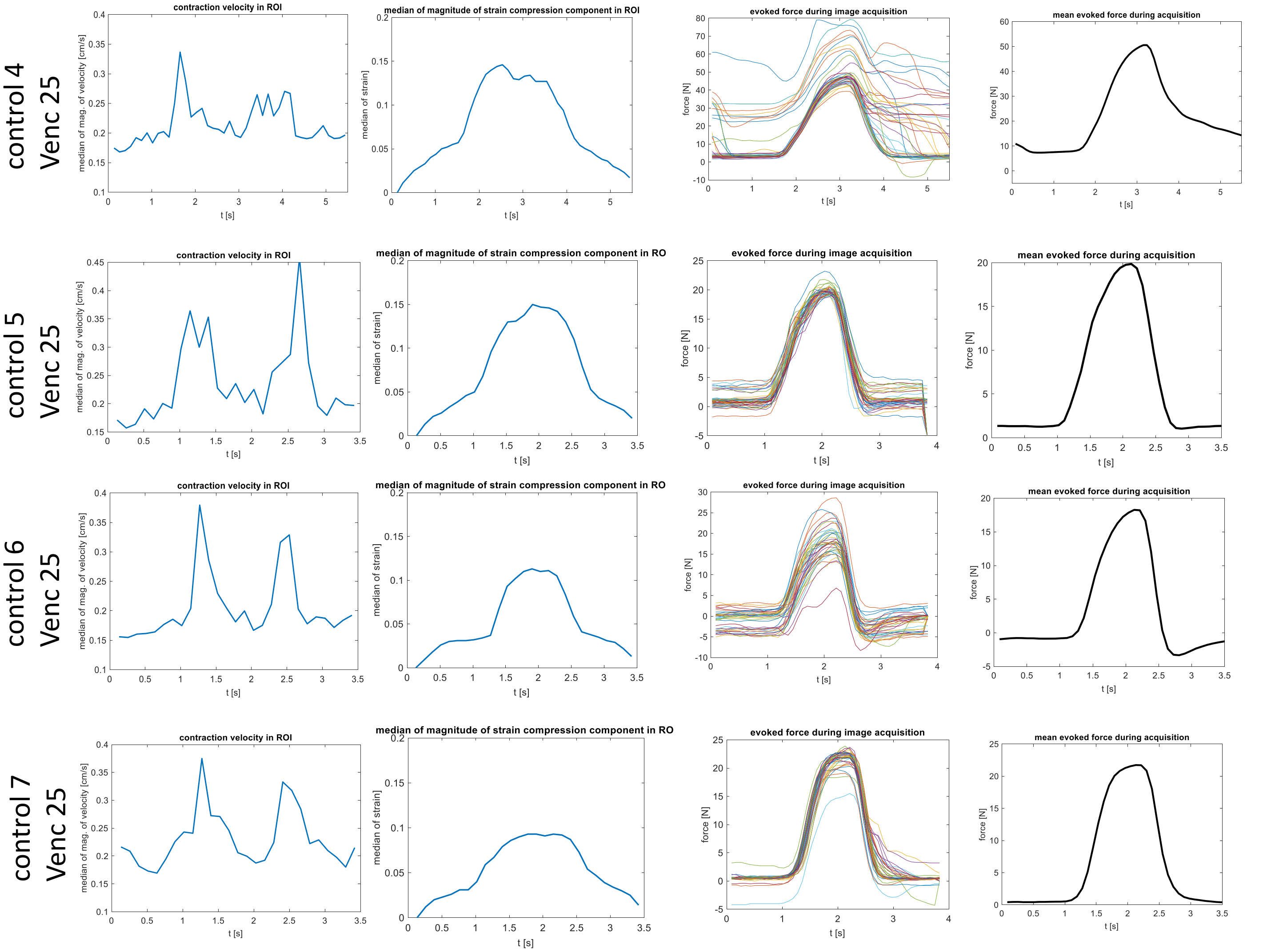

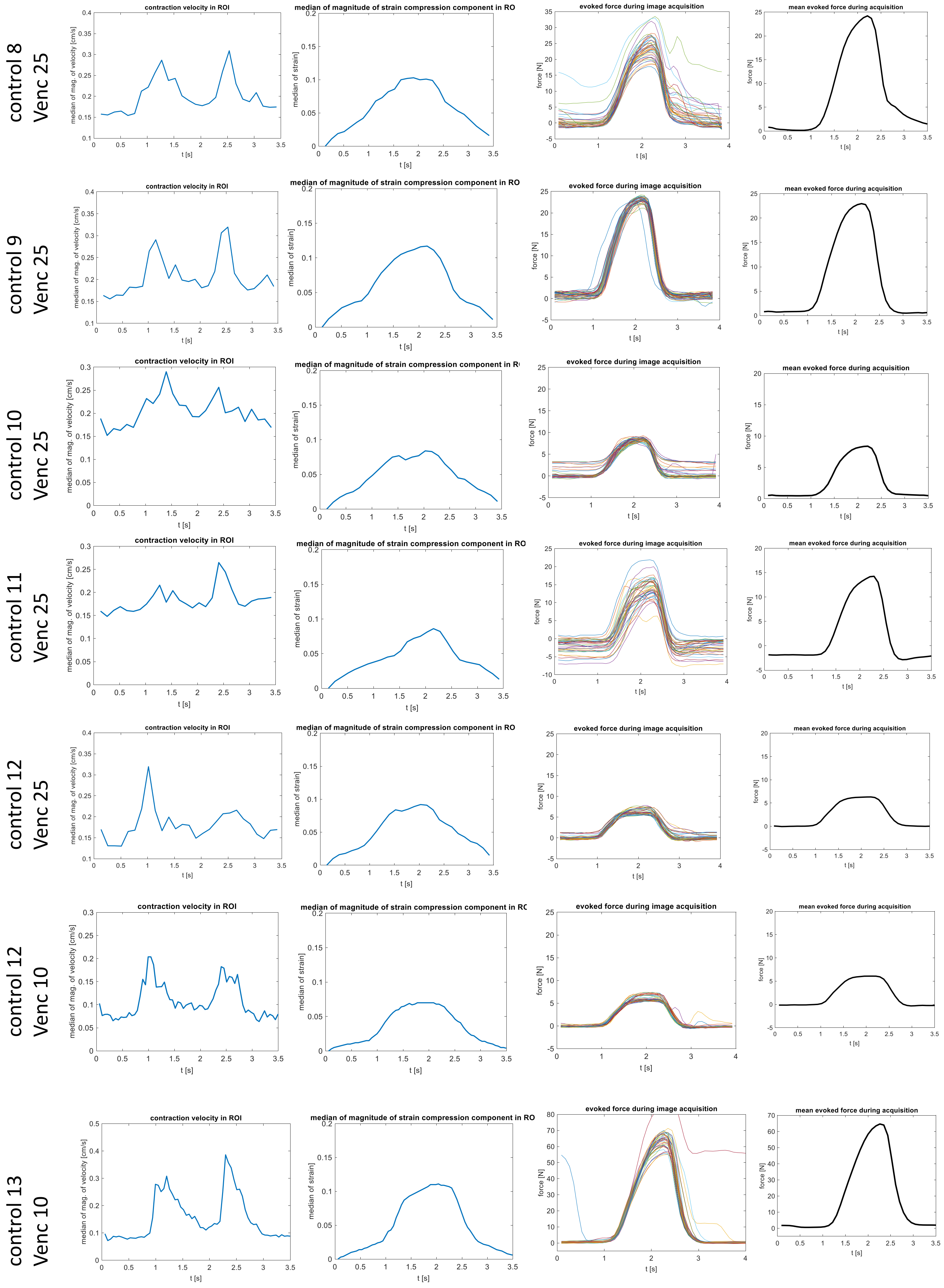
control 8
Venc 25
control 9
Venc 25
control 10
Venc 25
control 11
Venc 25
control 12
Venc 25
control 12
Venc 10
control 13
Venc 10
contraction velocity in ROI
median of magnitude of strain compression component in RO
evoked force during image acquisition
mean evoked force during acquisition
median of mag. of velocity [cm/s]
median of strain]
force [N]
t [s]

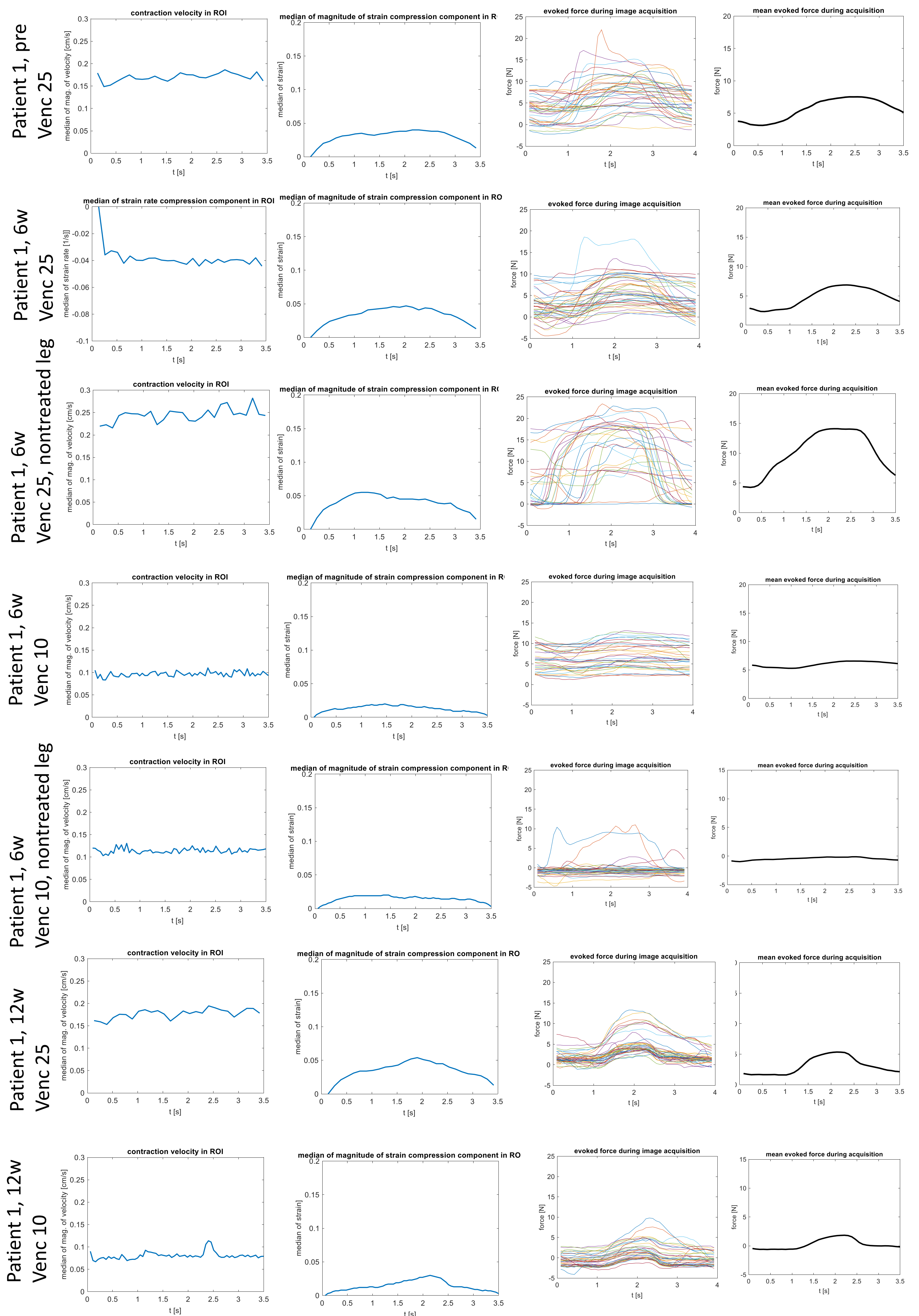

Patient 1, pre
Venc 25
contraction velocity in ROI
median of mag. of velocity [cm/s]
t [s]
median of magnitude of strain compression component in R
median of strain]
evoked force during image acquisition
force [N]
mean evoked force during acquisition
Patient 1, 6w
Venc 25
median of strain rate compression component in ROI
median of strain rate [1/s]]
median of magnitude of strain compression component in RO
Patient 1, 6w
Venc 25, nontreated leg
Patient 1, 6w
Venc 10
Patient 1, 6w
Venc 10, nontreated leg
Patient 1, 12w
Venc 25
Patient 1, 12w
Venc 10

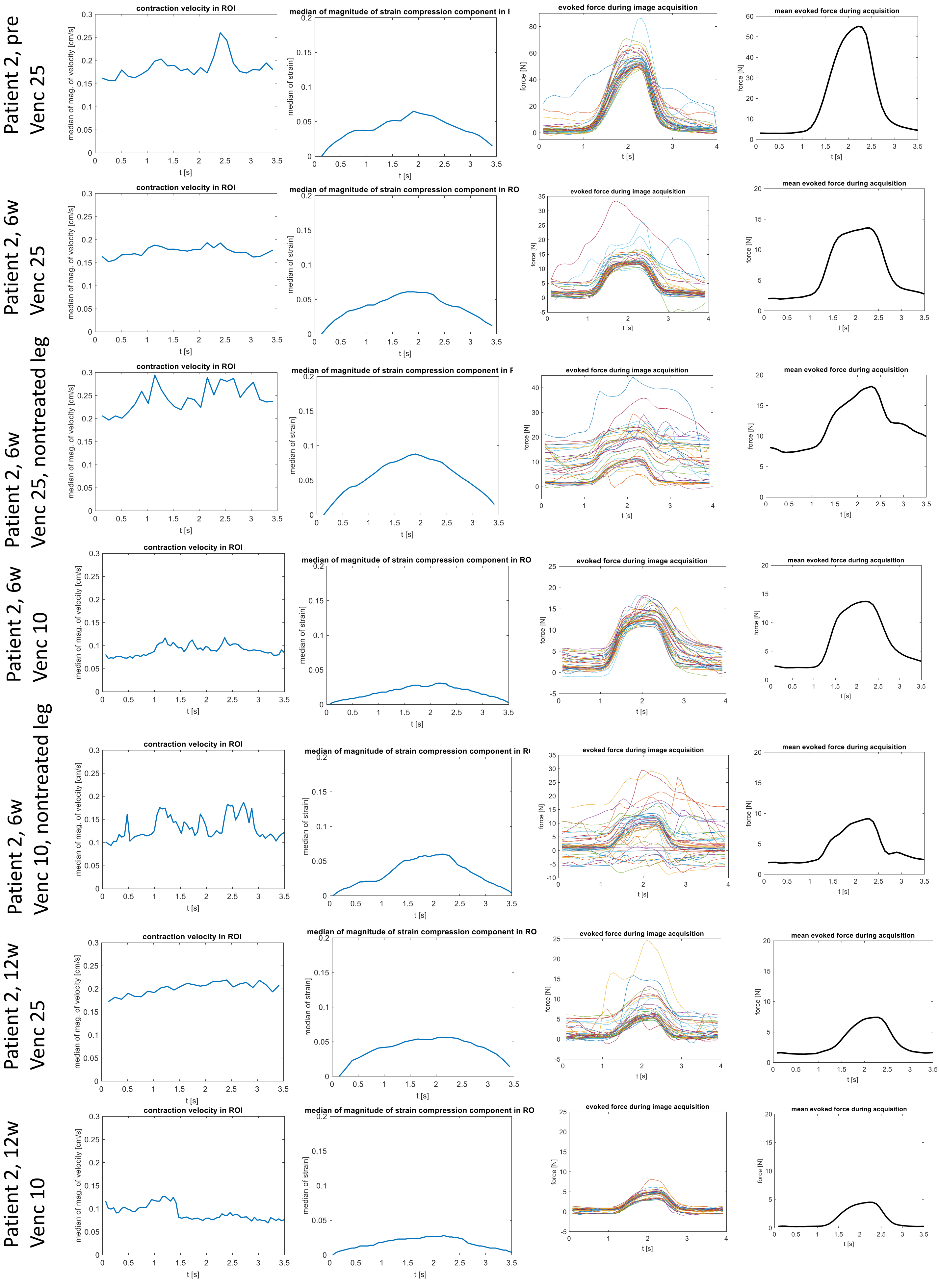

Patient 2, pre
Venc 25
Patient 2, 6w
Venc 25
Patient 2, 6w
Venc 25, nontreated leg
Patient 2, 6w
Venc 10
Patient 2, 6w
Venc 10, nontreated leg
Patient 2, 12w
Venc 25
Patient 2, 12w
Venc 10
contraction velocity in ROI
median of mag. of velocity [cm/s]
median of magnitude of strain compression component in ROI
median of strain]
evoked force during image acquisition
force [N]
mean evoked force during acquisition
t [s]

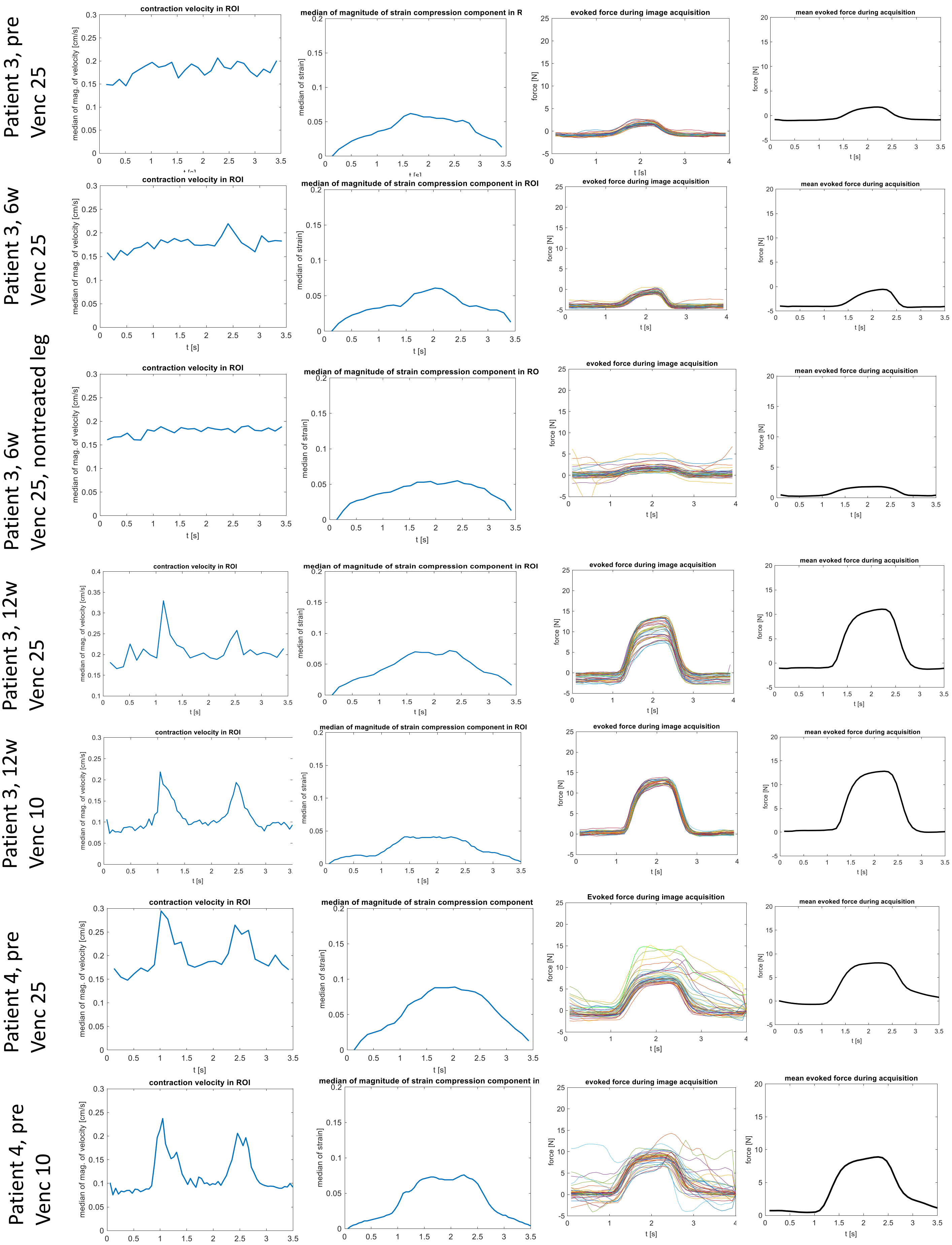

Patient 3, pre Venc 25
Patient 3, 6w Venc 25
Patient 3, 6w Venc 25, nontreated leg
Patient 3, 12w Venc 25
Patient 3, 12w Venc 10
Patient 4, pre Venc 25
Patient 4, pre Venc 10
contraction velocity in ROI
median of magnitude of strain compression component in ROI
evoked force during image acquisition
mean evoked force during acquisition
Evoked force during image acquisition
median of mag. of velocity [cm/s]
median of strain]
force [N]
t [s]

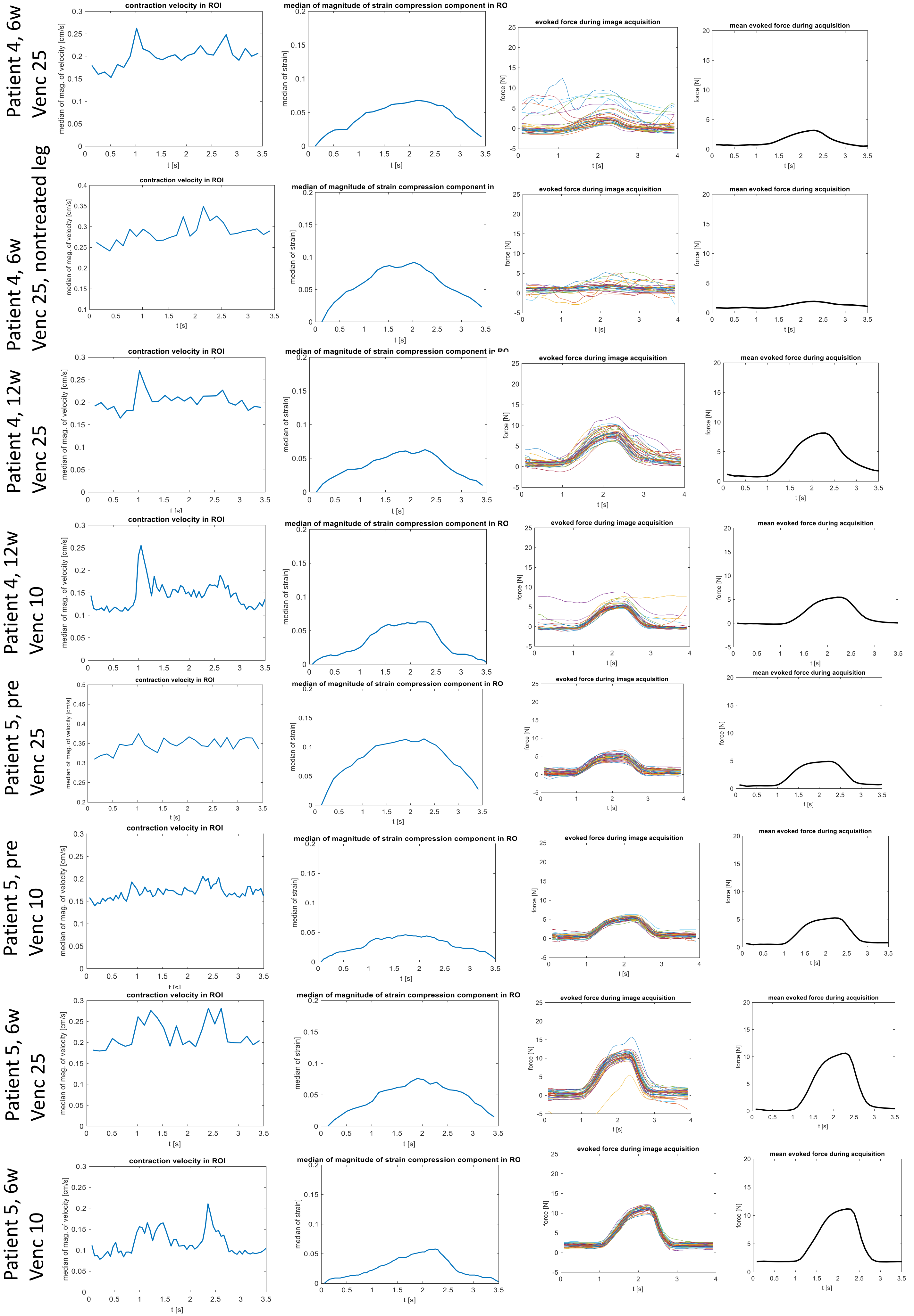
Patient 4, 6w
Venc 25
Patient 4, 6w
Venc 25, nontreated leg
Patient 4, 12w
Venc 25
Patient 4, 12w
Venc 10
Patient 5, pre
Venc 25
Patient 5, pre
Venc 10
Patient 5, 6w
Venc 25
Patient 5, 6w
Venc 10
contraction velocity in ROI
median of mag. of velocity [cm/s]
median of magnitude of strain compression component in RO
median of strain]
evoked force during image acquisition
mean evoked force during acquisition
force [N]
t [s]

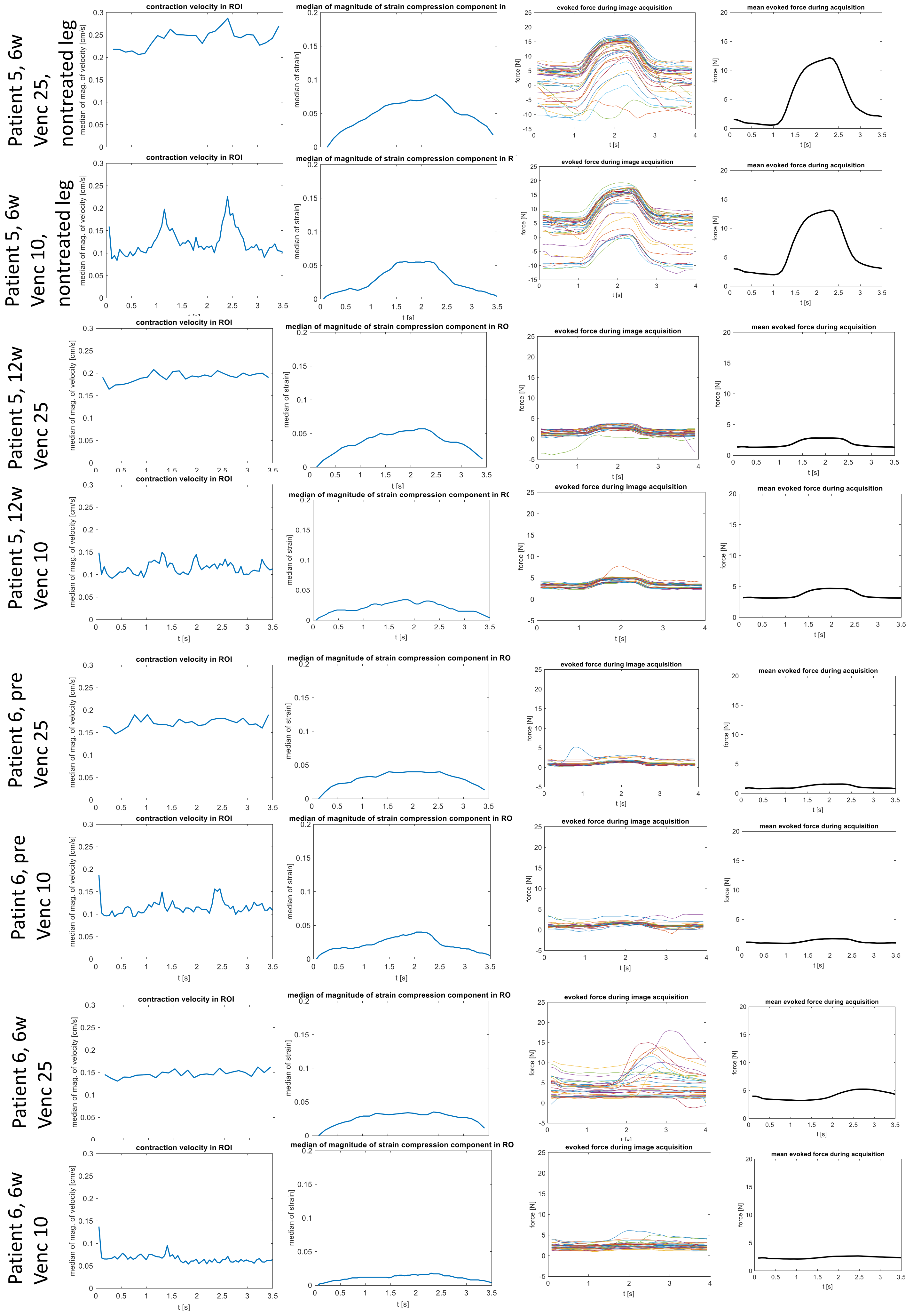
Patient 5, 6w
Venc 25,
nontreated leg
Patient 5, 6w
Venc 10,
nontreated leg
Patient 5, 12w
Venc 25
Patient 5, 12w
Venc 10
Patient 6, pre
Venc 25
Patint 6, pre
Venc 10
Patient 6, 6w
Venc 25
Patient 6, 6w
Venc 10
contraction velocity in ROI
median of mag. of velocity [cm/s]
median of magnitude of strain compression component in ROI
median of strain]
evoked force during image acquisition
force [N]
mean evoked force during acquisition
t [s]

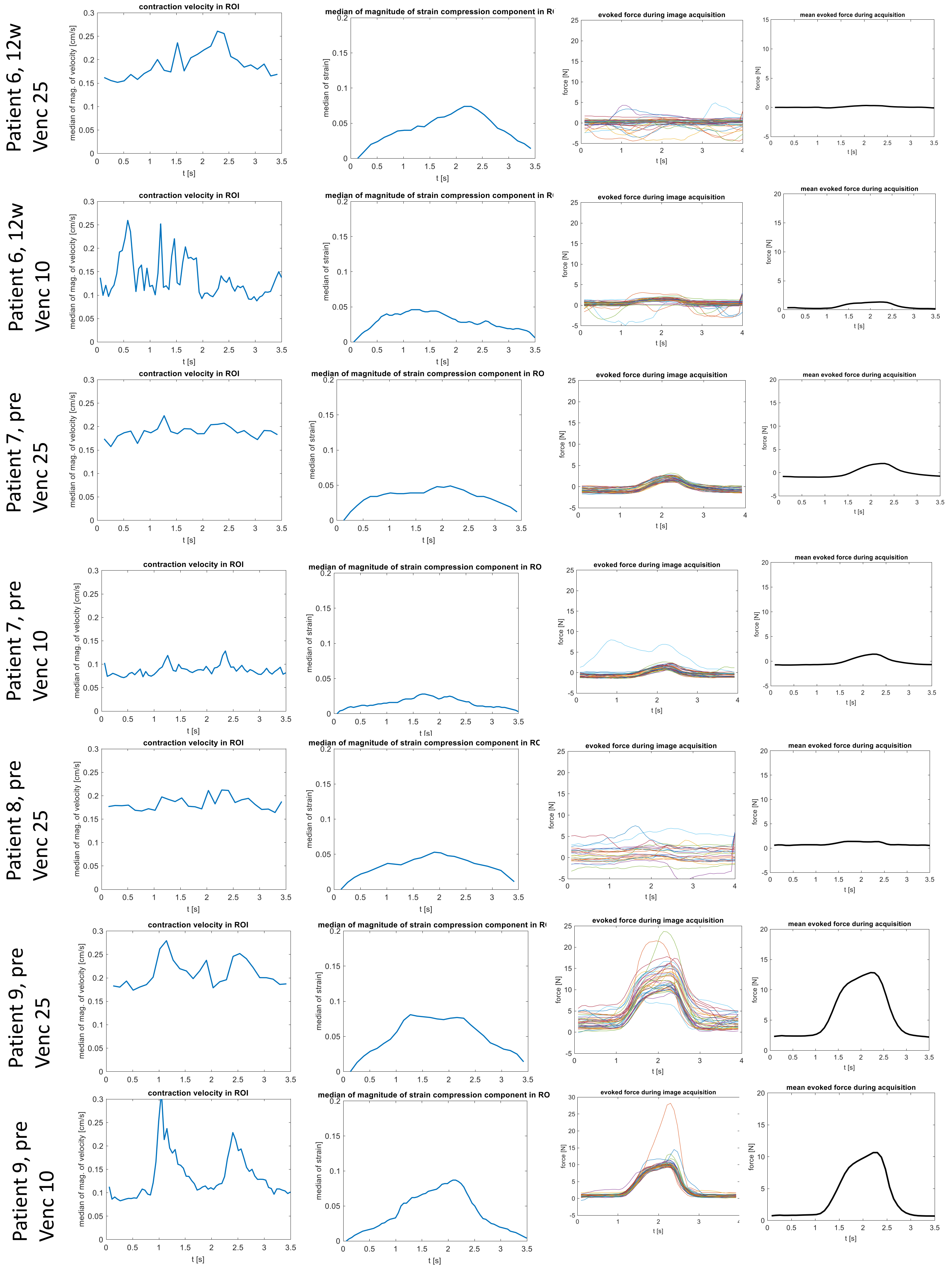

Patient 6, 12w Venc 25
Patient 6, 12w Venc 10
Patient 7, pre Venc 25
Patient 7, pre Venc 10
Patient 8, pre Venc 25
Patient 9, pre Venc 25
Patient 9, pre Venc 10
contraction velocity in ROI
median of mag. of velocity [cm/s]
median of magnitude of strain compression component in RO
median of strain]
evoked force during image acquisition
mean evoked force during acquisition
force [N]
t [s]

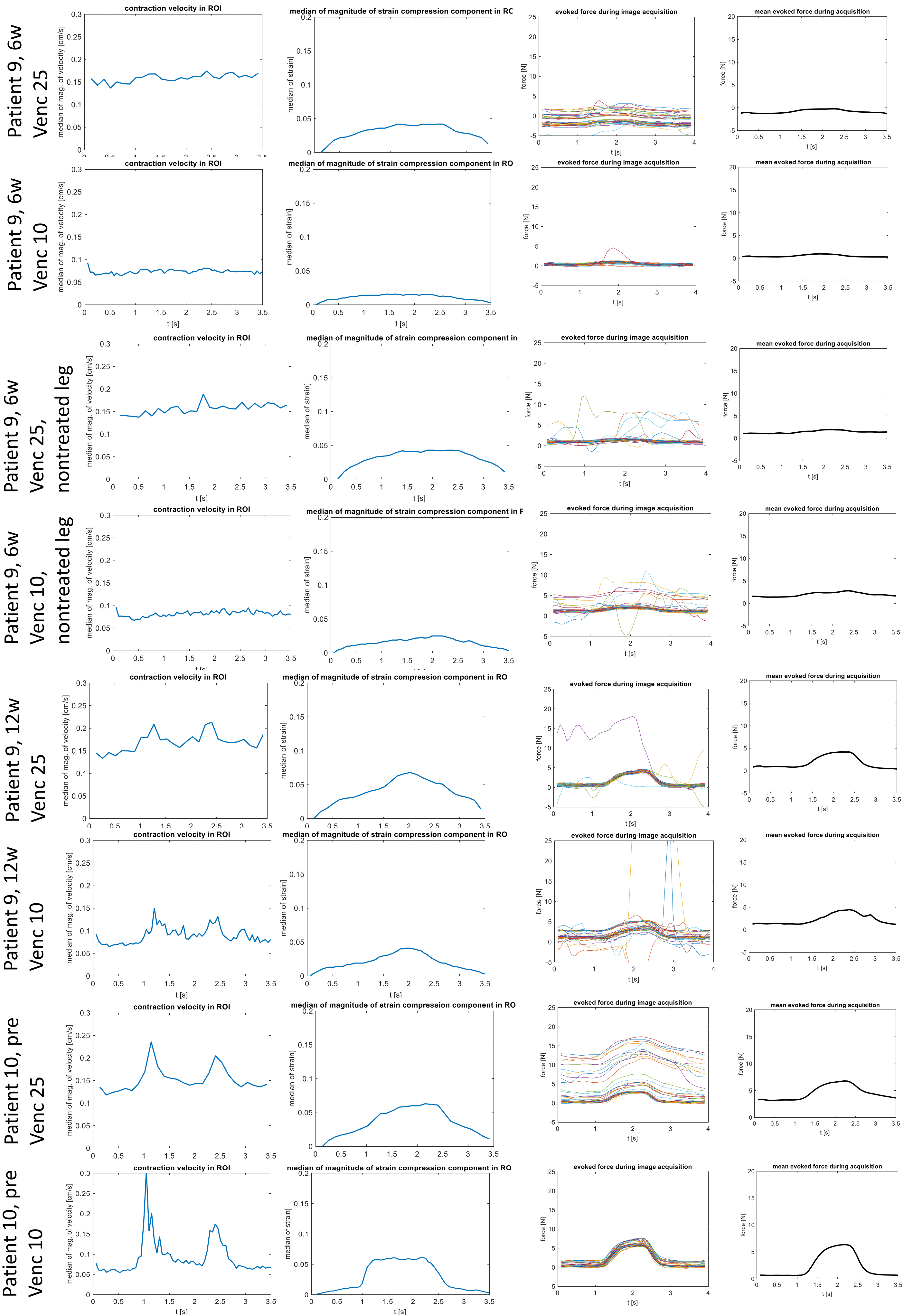
Patient 9, 6w
Venc 25
Patient 9, 6w
Venc 10
Patient 9, 6w
Venc 25,
nontreated leg
Patient 9, 6w
Venc 10,
nontreated leg
Patient 9, 12w
Venc 25
Patient 9, 12w
Venc 10
Patient 10, pre
Venc 25
Patient 10, pre
Venc 10
contraction velocity in ROI
median of mag. of velocity [cm/s]
median of magnitude of strain compression component in RO
median of strain]
evoked force during image acquisition
force [N]
mean evoked force during acquisition
t [s]

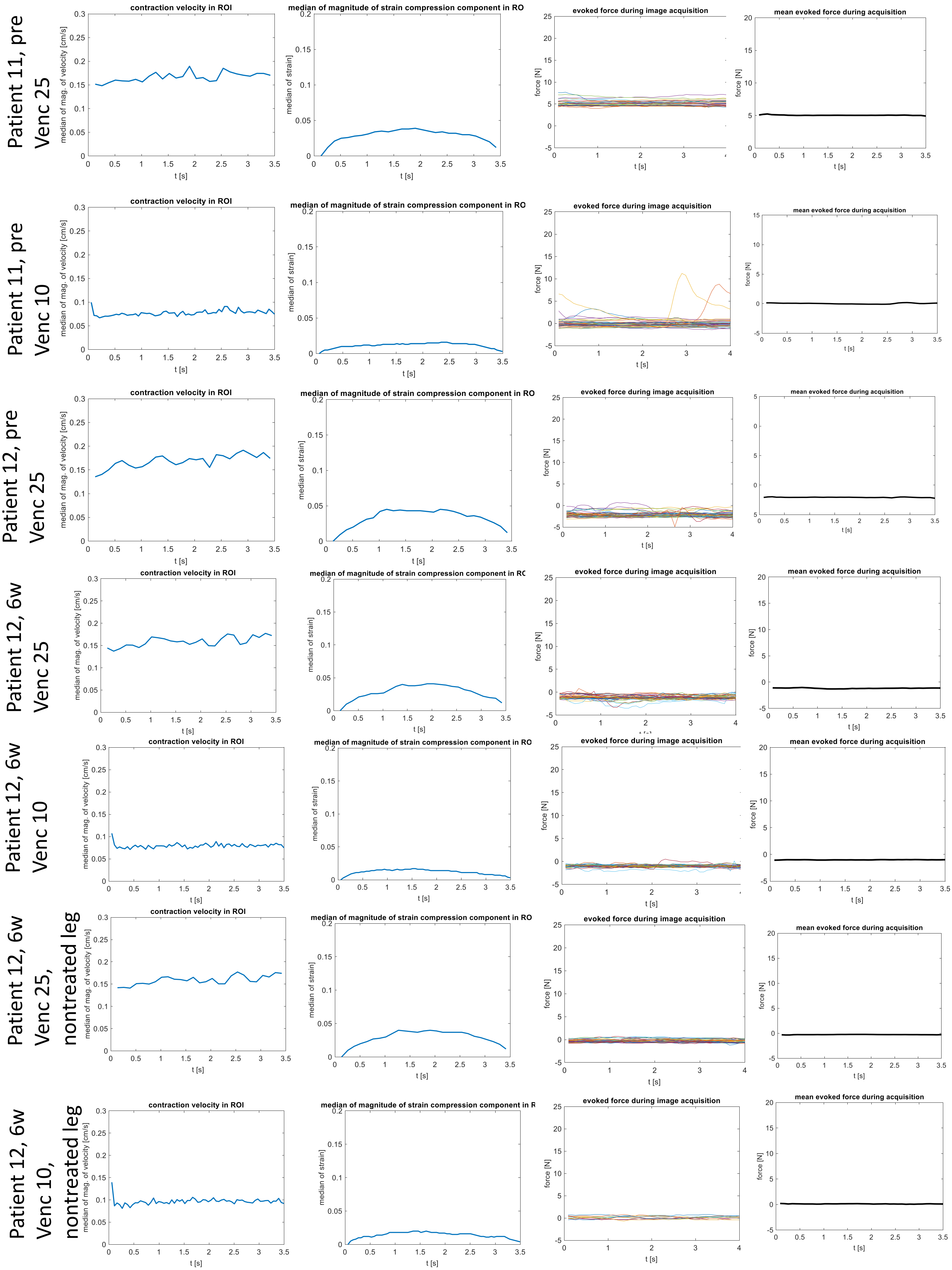

Patient 11, pre Venc 25
Patient 11, pre Venc 10
Patient 12, pre Venc 25
Patient 12, 6w Venc 25
Patient 12, 6w Venc 10
Patient 12, 6w Venc 25, nontreated leg
Patient 12, 6w Venc 10, nontreated leg
contraction velocity in ROI
median of mag. of velocity [cm/s]
median of magnitude of strain compression component in RO
median of strain]
evoked force during image acquisition
force [N]
mean evoked force during acquisition
t [s]

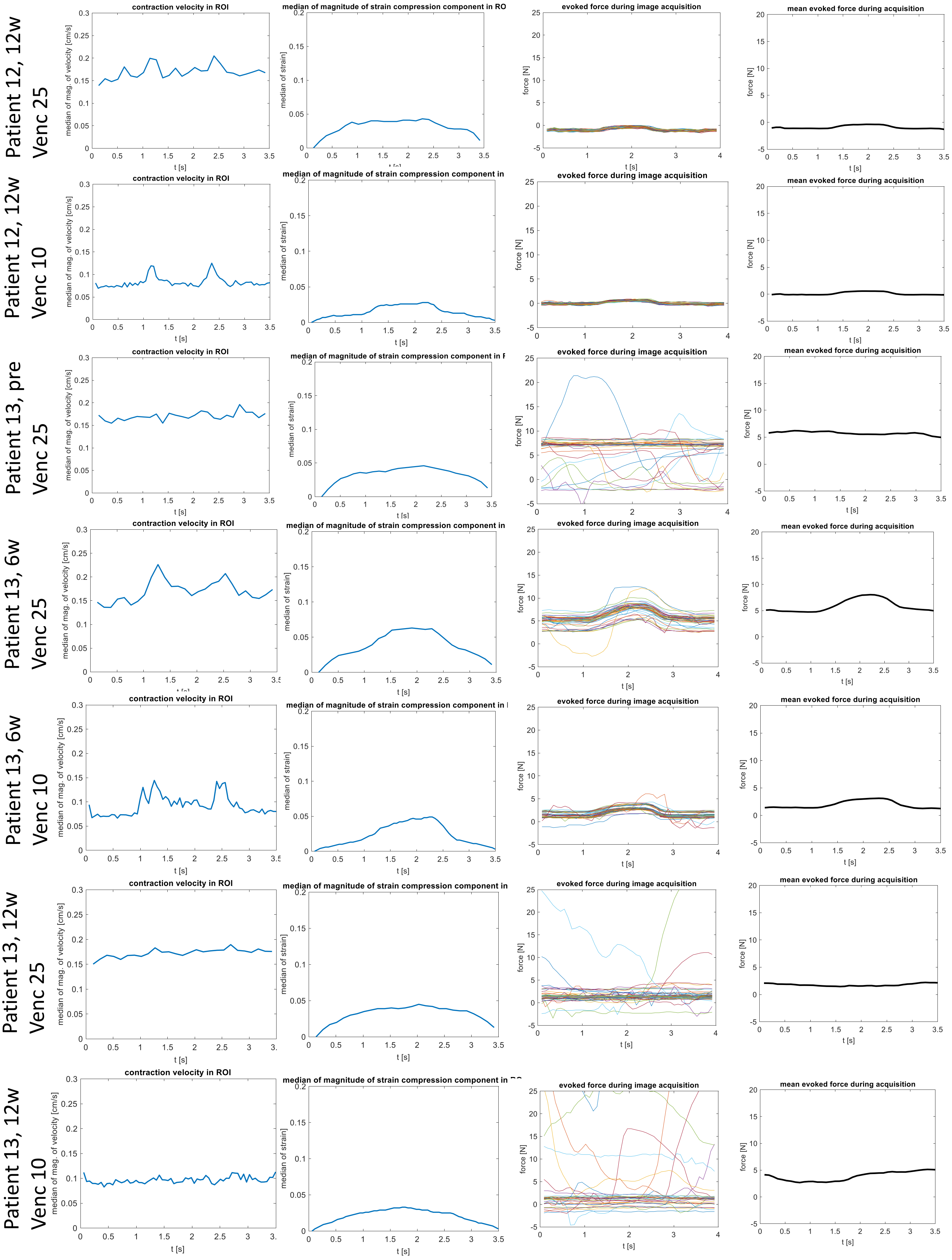
Patient 12, 12w Venc 25
Patient 12, 12w Venc 10
Patient 13, pre Venc 25
Patient 13, 6w Venc 25
Patient 13, 6w Venc 10
Patient 13, 12w Venc 25
Patient 13, 12w Venc 10
contraction velocity in ROI
median of mag. of velocity [cm/s]
median of magnitude of strain compression component in ROI
median of strain
evoked force during image acquisition
mean evoked force during acquisition
force [N]
t [s]

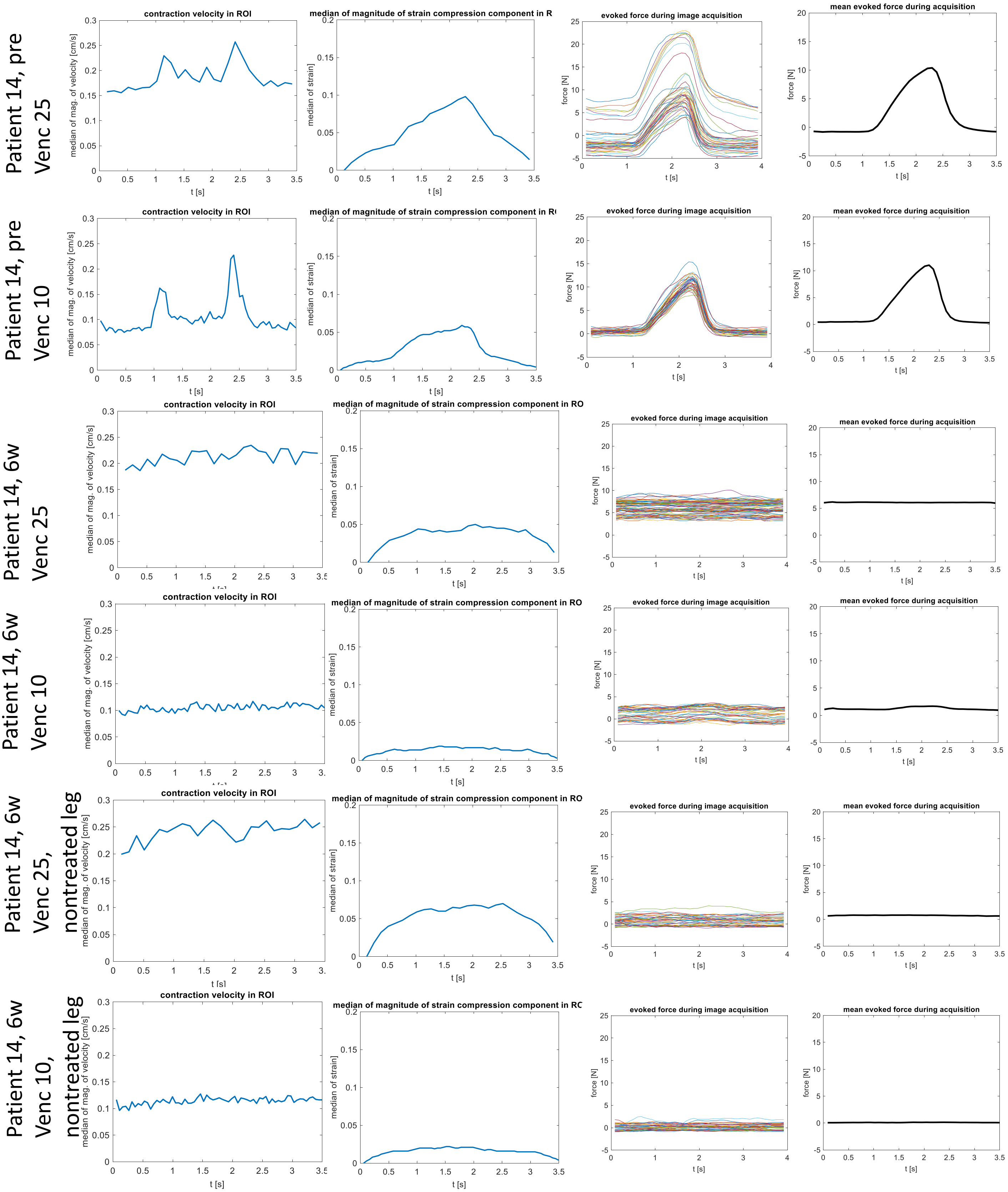
Patient 14, pre Venc 25
Patient 14, pre Venc 10
Patient 14, 6w Venc 25
Patient 14, 6w Venc 10
Patient 14, 6w Venc 25, nontreated leg
Patient 14, 6w Venc 10, nontreated leg
contraction velocity in ROI
median of mag. of velocity [cm/s]
median of magnitude of strain compression component in R
median of strain]
evoked force during image acquisition
force [N]
mean evoked force during acquisition
t [s]
