## Supplementary figures and images for "Cine phase contrast magnetic resonance imaging of calf muscle contraction in pediatric patients with cerebral palsy and healthy children: comparison of voluntary motion and electrically evoked motion"

### S2 Graphs

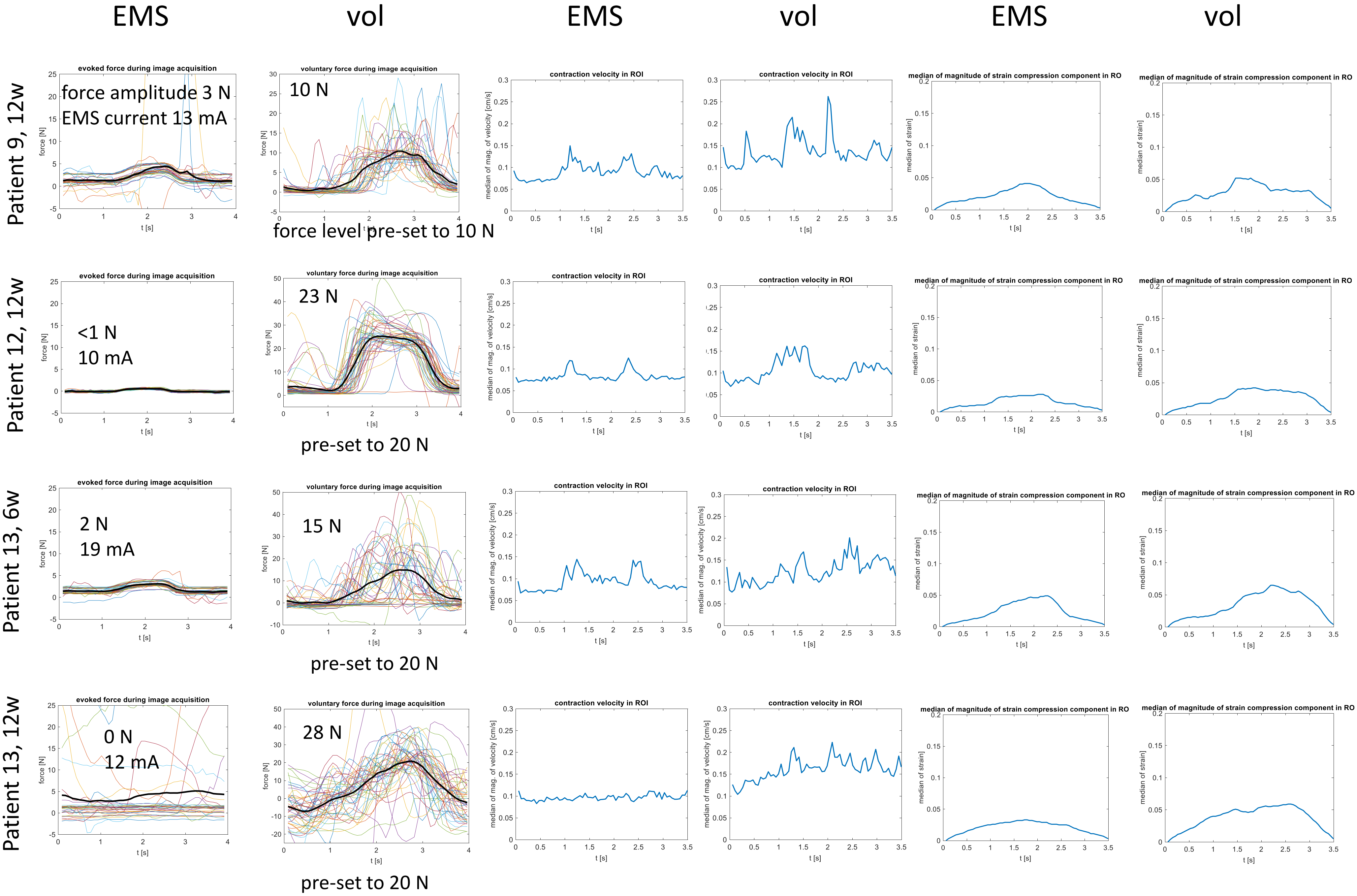

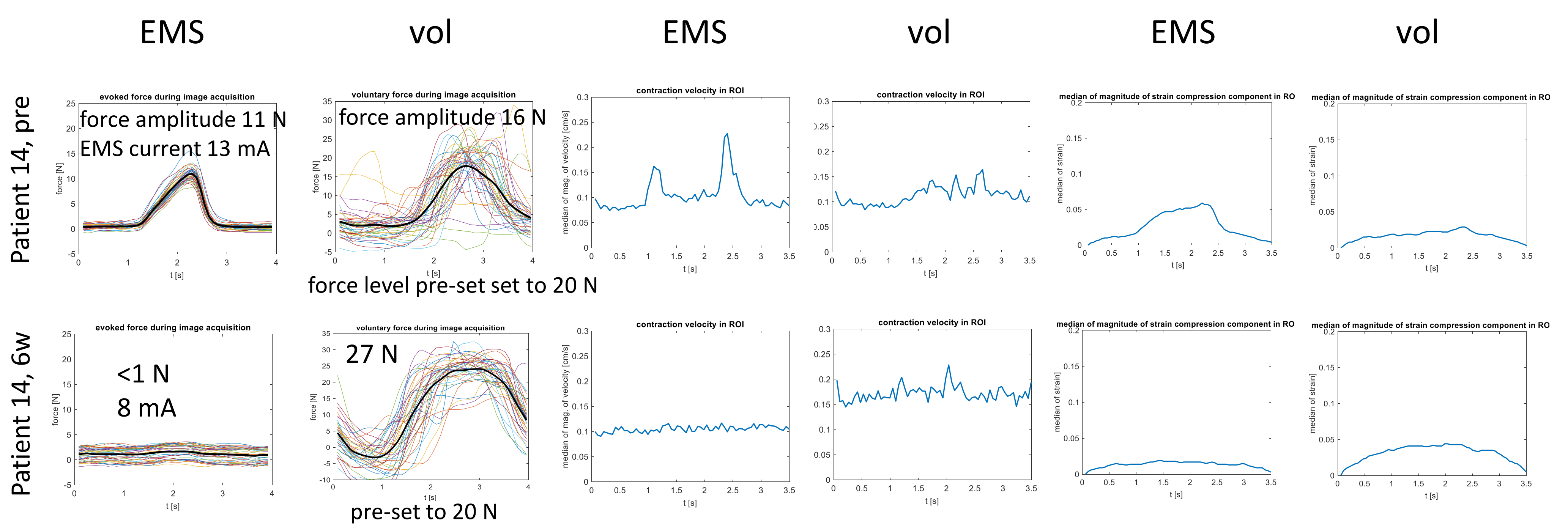

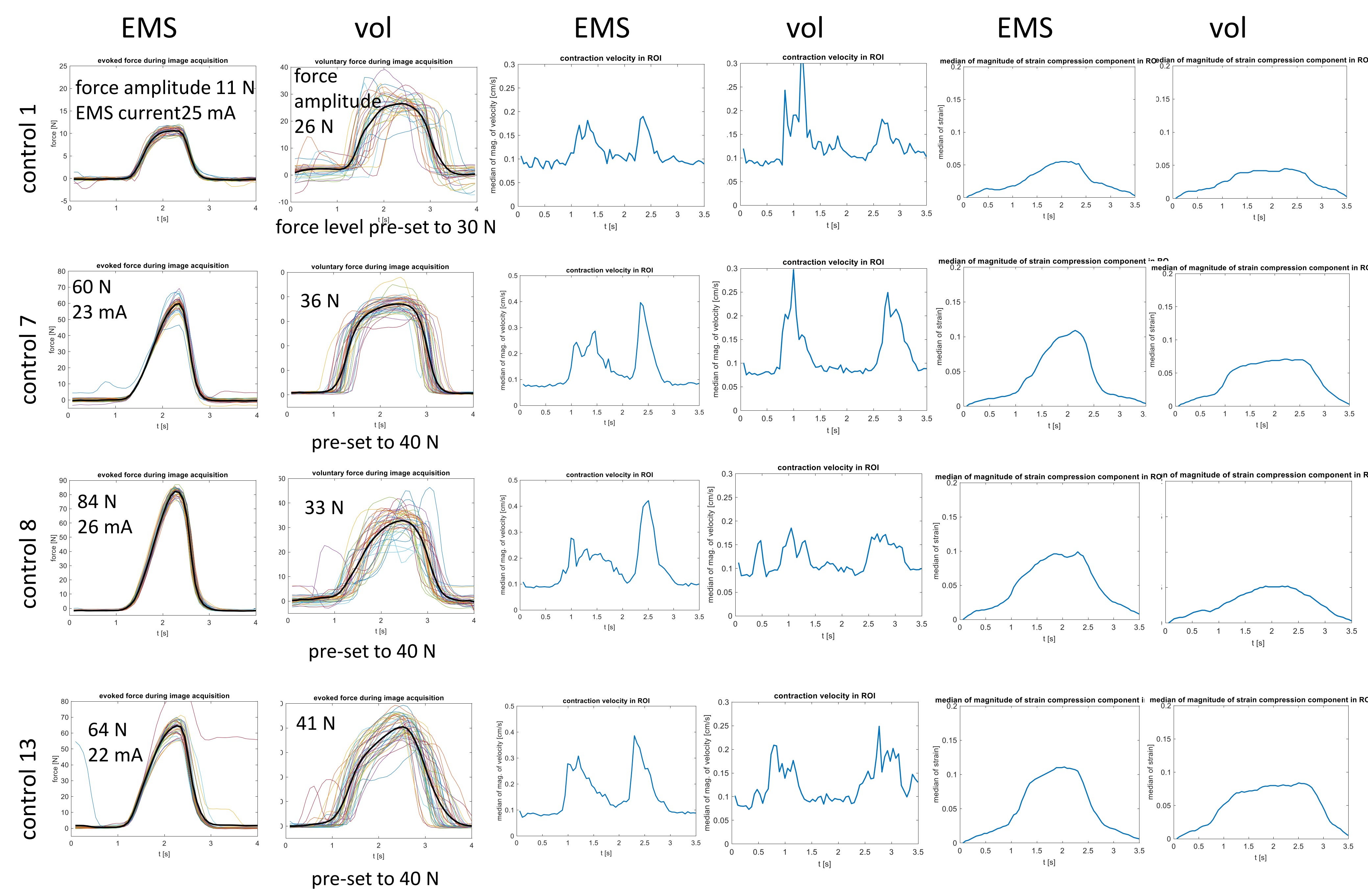
