## Supplementary material for "Cine phase contrast magnetic resonance imaging of calf muscle contraction in pediatric patients with cerebral palsy and healthy children: comparison of voluntary motion and electrically evoked motion": S3 Table

|  | max. voluntary force [N] | | | |
| --- | --- | --- | --- | --- |
| patient number | pre BTX | 6w post BTX | 6w post BTX, non-treated leg | 12w post BTX |
| 1 | 94 | 108 | 182 | 113 |
| 2 | 258 | 129 | 342 | 254 |
| 3 | 121 | 108 | 131 | 96 |
| 4 | 64 | 12 | 114 | 84 |
| 5 | 46 | 70 | 72 | 76 |
| 6 | 36 | 65 | 100 | 40 |
| 7 | 92 |  |  |  |
| 8 | 143 |  |  |  |
| 9 | 127 | 208 | 253 | 199 |
| 10 | 177 |  |  |  |
| 11 | 71 |  |  |  |
| 12 | 157 | 163 | 218 | 219 |
| 13 | 172 | 152 | 286 | 135 |
| 14 | 104 | 83 | 108 |  |
|  | max. voluntary force [N] | |  |  |
| control  number | 1st session | 2nd session |  |  |
| 1 | 275 | 273 |  |  |
| 2 | 388 |  |  |  |
| 3 | 326 |  |  |  |
| 4 | 292 |  |  |  |
| 5 | 257 |  |  |  |
| 6 | 260 |  |  |  |
| 7 | 151 | 273 |  |  |
| 8 | 273 | 268 |  |  |
| 9 | 155 |  |  |  |
| 10 | 170 |  |  |  |
| 11 | 108 |  |  |  |
| 12 | 207 |  |  |  |
| 13 | 343 |  |  |  |
